# Audited large language model triage for systematic review screening in national clinical guideline production: validation and prospective deployment

**DOI:** 10.64898/2026.06.02.26354724

**Authors:** Petter Fagerberg, Oscar Sallander, Kim Vikhe Patil, Charlotta Thunborg, Lina Lundström, Anders Berg, Anastasia Nyman, Natalia Borg, Thomas Lindén

## Abstract

Title and abstract screening limit the timeliness of systematic reviews used for clinical guidelines. We evaluated audited large language model (LLM) triage at Sweden’s National Board of Health and Welfare. Ten LLMs from five model families were tested on 419 Cochrane reviews comprising 26,892 records, and the selected ensemble was externally validated on 133 reviews including 8,501 records matched to planned guideline topics. The same locked model pair was then used prospectively across 24 systematic reviews in two national guideline programmes. On the 419-review selection benchmark, the selected Gemini-3-flash plus GPT-5.1 ensemble achieved 98.0% (95% CI, 97.3–98.7) mean review-level sensitivity, while topic-matched validation yielded 96.7% sensitivity (95% CI, 93.7–98.9). Prospective deployment screened 74,679 records, placed 63,858 (85.5%) in the AI-excluded pool and reduced estimated first-pass screening effort from 415 to 34 person-days. Across 600 randomly sampled AI-excluded records from the migraine and dementia programmes, none was confirmed as a final false negative after post-unblinding adjudication; across the completed 680-record audit, all 38 final retained records had been AI flagged, whereas locked blinded human consensus missed seven. These findings support locked, audited LLM triage, with human oversight and programme-specific monitoring, for systematic reviews used in national guidelines.

## Introduction

Clinical guidelines shape day-to-day medical practice and depend on systematic reviews, yet their timeliness is constrained by a persistent bottleneck: title and abstract screening. This step consumes substantial expert time before evidence appraisal, recommendation development or guideline updating can begin^1–4^.

Current screening policies force a trade-off between workload and redundancy. Dual human independent screening with consensus remains the high-resource reference standard but is difficult to sustain when searches return tens of thousands of records across multiple guideline questions. Single-reviewer screening reduces workload, but leaves exclusion decisions more vulnerable to fatigue and individual error; empirical screening studies and methodological reviews show that single screening can miss relevant studies, and that duplicate screening reduces but does not eliminate this risk^5–7^. An audited AI approach could be a credible alternative to the manual human process only if it is selected, validated and deployed under explicit governance, and evaluated against these realistic human alternatives rather than an idealised baseline^8^. For guideline producers, AI-assisted screening cannot be judged by classification accuracy alone. It must also preserve transparency, accountability and the ability to recover studies that could change a review’s conclusions^9,10^.

Earlier machine learning automation tools showed that screening workload can be reduced. For example, Chan et al.^11^ provided a strong benchmark dataset^12^ and trained a specialized BioBERT screening model using around 540,000 labelled title and abstract decisions from the Cochrane library corpus. While the model enabled high sensitivity screening at scale, its reliance on a very large in-domain training corpus limits its generalisability, particularly for under-represented topics in which false-negative rates were high^11^. This leaves uncertainty about how well such supervised models translate to real-world guideline production, where topics may fall outside Cochrane’s corpus, and adoption of such tools has been limited^2,13,14^. In contrast, contemporary LLMs are pre-trained on broad, heterogeneous corpora and may therefore offer greater flexibility across review topics, since they can often be adapted through prompting rather than topic-specific labelled training data. More recently, LLM studies report high sensitivity and workload reduction across prompt templates, compact models, multi-model ensembling and multi-agent designs^15–21^, but prospective evidence for deployed medical AI systems remains a central concern^10^. In evidence synthesis specifically, Cochrane has published a protocol for evaluating semi-automated review methods across title and abstract screening, full-text screening and data extraction^8^. However, prospective deployment evidence in live guideline production remains limited^22^.

In earlier work we showed that state-of-the-art LLMs can screen titles and abstracts in batches of 100 without sensitivity loss^20^, and that a dual-model ensemble with two runs per model can increase sensitivity while retaining acceptable specificity^16^. Those studies did not answer how model choice should be validated across rapid successive LLM model releases, whether performance generalises to target guideline domains, how workload reduction translates to operational corpora, or how such a process should be governed prospectively in national guideline production.

Here we report a staged validation to deployment study of audited LLM triage inside Sweden’s National Board of Health and Welfare, the country’s statutory guideline-producing agency^23^. We benchmarked ten LLMs across five model families on 419 Cochrane reviews, externally validated the selected ensemble on 133 reviews matched to the prospective guideline topics, deployed it prospectively across 24 systematic reviews in two national guidelines under locked models and prespecified audit, and compared its performance against realistic dual and single human screening alternatives. To our knowledge, this is the first prospective deployment study of audited LLM triage inside a national guideline-producing agency.

## Results

### Retrospective validation across 419 Cochrane reviews

Ten LLMs across five model families were evaluated on 26,892 screening records (4,545 final inclusions) from 419 Cochrane reviews spanning 53 Cochrane review groups (Fig. 1a; Extended Data Fig. 1; Extended Data Table 1, Supplementary Table 9). On this 419-SR selection benchmark, the selected closed-source dual-model dual-run ensemble, Gemini-3-flash plus GPT-5.1, achieved a mean review-level sensitivity of 98.0% (95% review-clustered bootstrap interval, 97.3-98.7) across 361 reviews containing at least one included study (Extended Data Fig. 4; Supplementary Table 8). In total, the selected ensemble missed 82 final inclusions pooled across reviews (Extended Data Fig. 6). No included records were missed in 305 of 361 reviews (Extended Data Fig. 6).

**Fig. 1.**
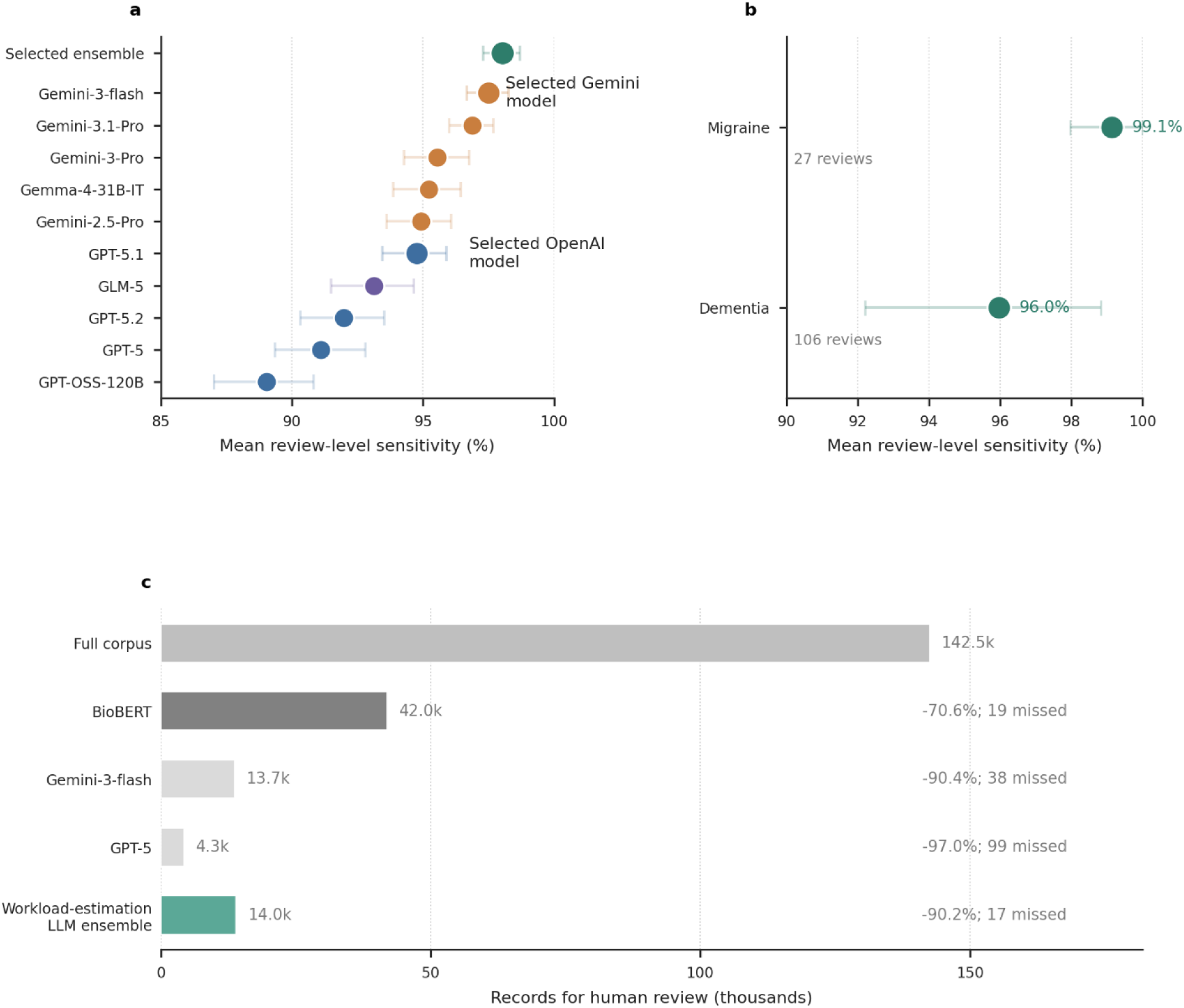
Validation, external generalisation and workload impact of the selected LLM ensemble. a, Model selection on the 419-SR validation set. Points show mean review-level sensitivity for the ten individual candidate LLMs and the selected ensemble. Horizontal error bars show 95% review-clustered bootstrap intervals. The selected ensemble combined the highest-sensitivity Gemini model (Gemini-3-flash), with the highest-sensitivity OpenAI model (GPT-5.1). Colours indicate model family; the selected ensemble is shown in green. b, External validation of the selected ensemble in guideline-related Cochrane review sets. Points show mean review-level sensitivity in migraine and dementia reviews matched to the target national guideline programmes. Horizontal bars show 95% review-clustered bootstrap intervals. c, Estimated workload impact in the 22-SR real-search workload. Bars show operational record counts routed to human review under manual screening, BioBERT (published Chan et al. visual comparator), individual LLM baselines and the closely related workload-estimation LLM ensemble using Gemini-3-flash plus GPT-5. Right-side annotations show workload reduction relative to manual screening and the number of missed included studies. Panels a and b report review-level performance estimates; panel c reports operational workload counts. Macro-mean review-level sensitivity is computed over the 25 of 27 migraine reviews and the 87 of 106 dementia reviews containing at least one final-included record.

The main open-weight comparator, Gemma-4-31B-IT plus GPT-OSS-120B, achieved a mean review-level sensitivity of 95.9% (94.7–97.0) on the 419-review validation set (Extended Data Fig. 4; Supplementary Table 8). Model provider family had the largest sensitivity impact (3.8 percentage points; 95% CI, 3.1–4.5), exceeding OR-ensembling (1.8 pp; 95% CI, 1.5–2.1) and cross-family pairing beyond the best single model (0.7 pp; 95% CI, 0.4–1.0; Extended Data Fig. 2).

Within-family model release-pair comparisons showed non-monotonic sensitivity and specificity changes across temporally ordered Gemini Pro plus GPT pairs (Extended Data Fig. 5; Supplementary Table 13). In the most recent transition (Gemini-3-Pro plus GPT-5.1 to Gemini-3.1-Pro plus GPT-5.2), the newer pair newly missed 43 included records while rescuing 31, despite very similar mean review-level sensitivity, supporting explicit revalidation rather than automatic upgrade on new model releases. The unanimous-exclusion label-discordance rate fluctuated between 0.49% and 0.96% across release pairs. Cross-family ensembling buffered but did not eliminate these changes.

### Topic-matched external validation

On 133 non-overlapping Cochrane reviews aligned with the prospective guideline domains, 27 migraine reviews comprising 1,343 records and 106 dementia reviews comprising 7,158 records, the selected ensemble achieved a mean review-level sensitivity of 96.7% (95% review-clustered bootstrap interval, 93.7–98.9), consistent with generalisability to the deployment domains (Fig. 1b; Extended Data Fig. 3; Supplementary Table 10). 25 out of 964 final inclusion records from Cochrane reference lists were missed across these reviews.

### Real-search workload reduction exceeded 90%

On the 22-review real-search corpus (142,504 records; 768 included), a closely related workload-estimation ensemble using the same Gemini component and the prior GPT generation (Gemini-3-flash plus GPT-5; substituted because the run completed before GPT-5.1 release) reduced the human review queue from 142,504 to 13,968 records, a pooled record-level workload reduction of 90.2% (95% review-clustered bootstrap interval, 85.7-92.7). The same operational pooled record-level analysis yielded sensitivity of 97.8% (95.7-99.4) and specificity of 90.7% (86.4-93.1), with 17 of 768 final Cochrane reference-list inclusion records missed across reviews; the corresponding mean review-level sensitivity and specificity were 98.3% (96.8-99.5) and 88.5% (84.5-92.0), respectively (Supplementary Table 10; Fig. 1c). Chan et al.’s published BioBERT comparator results, included here as an external visual benchmark rather than results generated in this study, achieved 97.5% pooled record-level sensitivity and 70.9% pooled record-level specificity on the same corpus11, routing approximately 28,000 more false-positive records to human review than the workload-estimation LLM ensemble, or approximately 156 additional reviewer-days under dual screening, while missing 19 final inclusion records.

### Prospective deployment in two national guidelines

The selected dual-model LLM ensemble, Gemini-3-flash plus GPT-5.1, was integrated into 24 prospective reviews in the Swedish national guidelines for migraine and dementia (Supplementary Table 5). The reviews spanned pharmacological, non-pharmacological, diagnostic, organisational, safety and competence-training questions, covering acute drug therapy, preventive drug therapy, neuromodulation devices, behavioural and psychological interventions, care pathways, diagnostic procedures, and clinical competence development across both guideline programmes (Supplementary Data Tables 6 and 7). Searches were executed programmatically across six database sources, deduplicated, and passed through the same four-run LLM-ensemble triage process. Across both programmes, 63,858 of 74,679 records (85.5%) received unanimous exclusion across all four runs and entered the AI-excluded pool; 10,821 records (14.5%) were routed to human review (Extended Data Fig. 7) on the basis of any non-unanimous-exclude signal. A prespecified audit then gave 680 sampled records additional dual-audit review (340 per guideline; 300 from the AI-excluded stratum and 40 from the AI-flagged stratum). Because the 80 AI-flagged audit records were already in the routine human-review stream, the resulting 11,501 value in Fig. 2 is a reviewer-facing workload count rather than a count of distinct records.

**Fig. 2.**
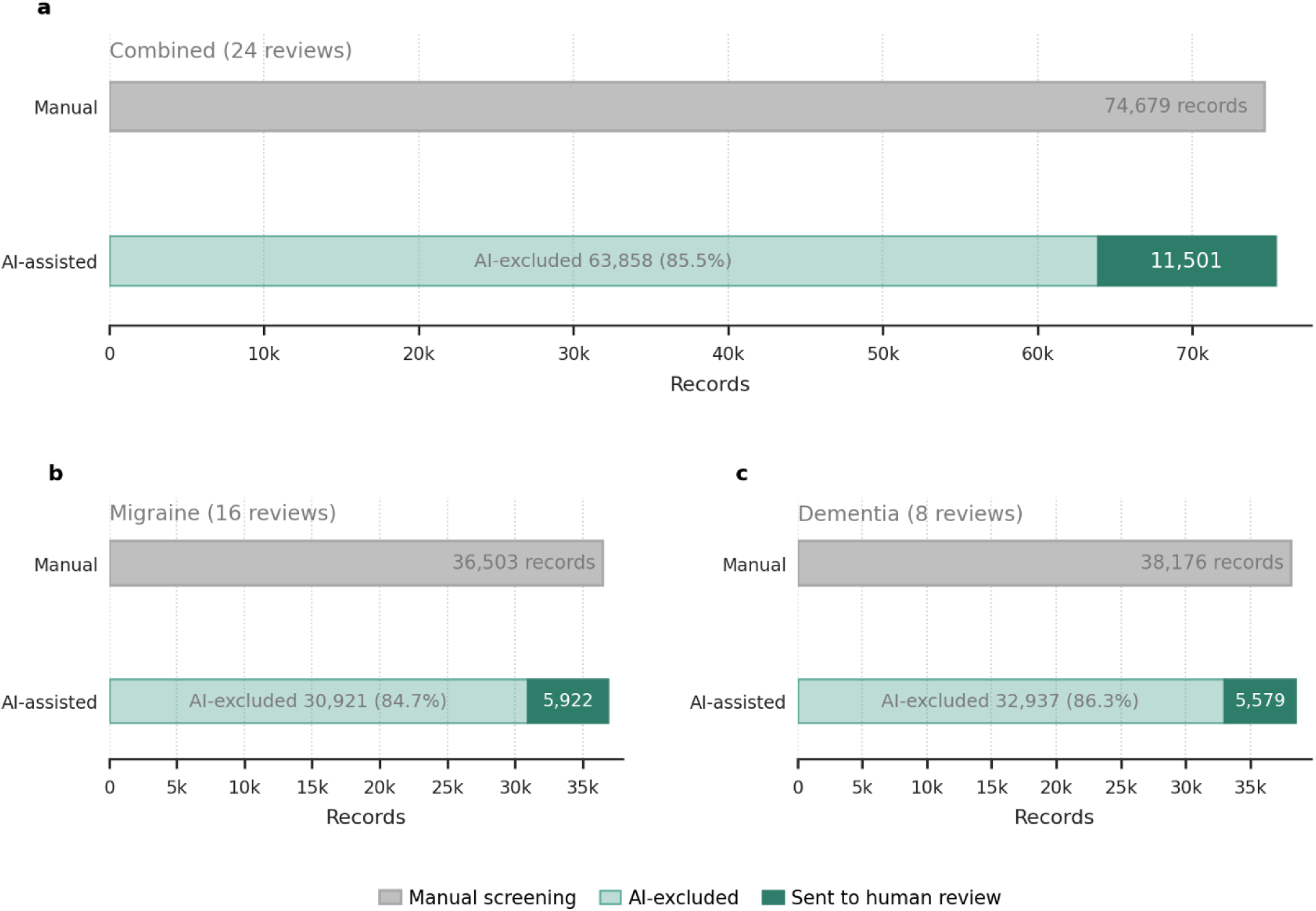
Prospective records requiring human review under manual and AI-assisted screening. a, Pooled record-level deployment summary across both national-guideline programmes (24 reviews, 74,679 records). All quantities shown are pooled operational counts or reviewer-facing workload counts, not inferential performance estimates. The AI ensemble routed 10,821 records (14.5%) to human review if any run returned a non-exclude decision. The prespecified audit gave 680 sampled records additional dual-audit review (340 per programme; 300 from the AI-excluded stratum and 40 from the AI-flagged stratum). Because 80 AI-flagged audit records were already in the routine human-review stream, the combined 11,501 value is a reviewer-facing workload count rather than a distinct-record count. b, c, Migraine and dementia deployments separately, each including a 340-record audit. Grey bars show the full manual-screening corpus. Green bars show the AI-assisted process, including AI-flagged records and the audit sample. Audit records are counted both within their originating pool and within the reviewer-facing workload count; consequently the green bar segments sum to 680 records more than the corpus total in panel a and 340 records more in panels b and c.

The screening process reduced estimated first-pass effort from 415 person-days under conventional dual human screening to 34 person-days under the deployed AI-assisted process under the base-case assumption of one minute per title and abstract decision and six screening hours per reviewer-day (Table 1). The corresponding sensitivity range was 207 to 17 person-days at 30 seconds per record and 830 to 68 person-days at two minutes per record, before adding implementation, adjudication-meeting and pipeline-maintenance overheads. The audit at the deployed sampling (300 AI-excluded records adjudicated per programme) excludes a programme-level false-negative rate above 1.0% in each AI-excluded stratum, and above 0.5% in the pooled 600-record AI-excluded audit (one-sided exact 95% upper bound, Clopper-Pearson), but does not exclude review-local misses below those rates.

**Table 1.**
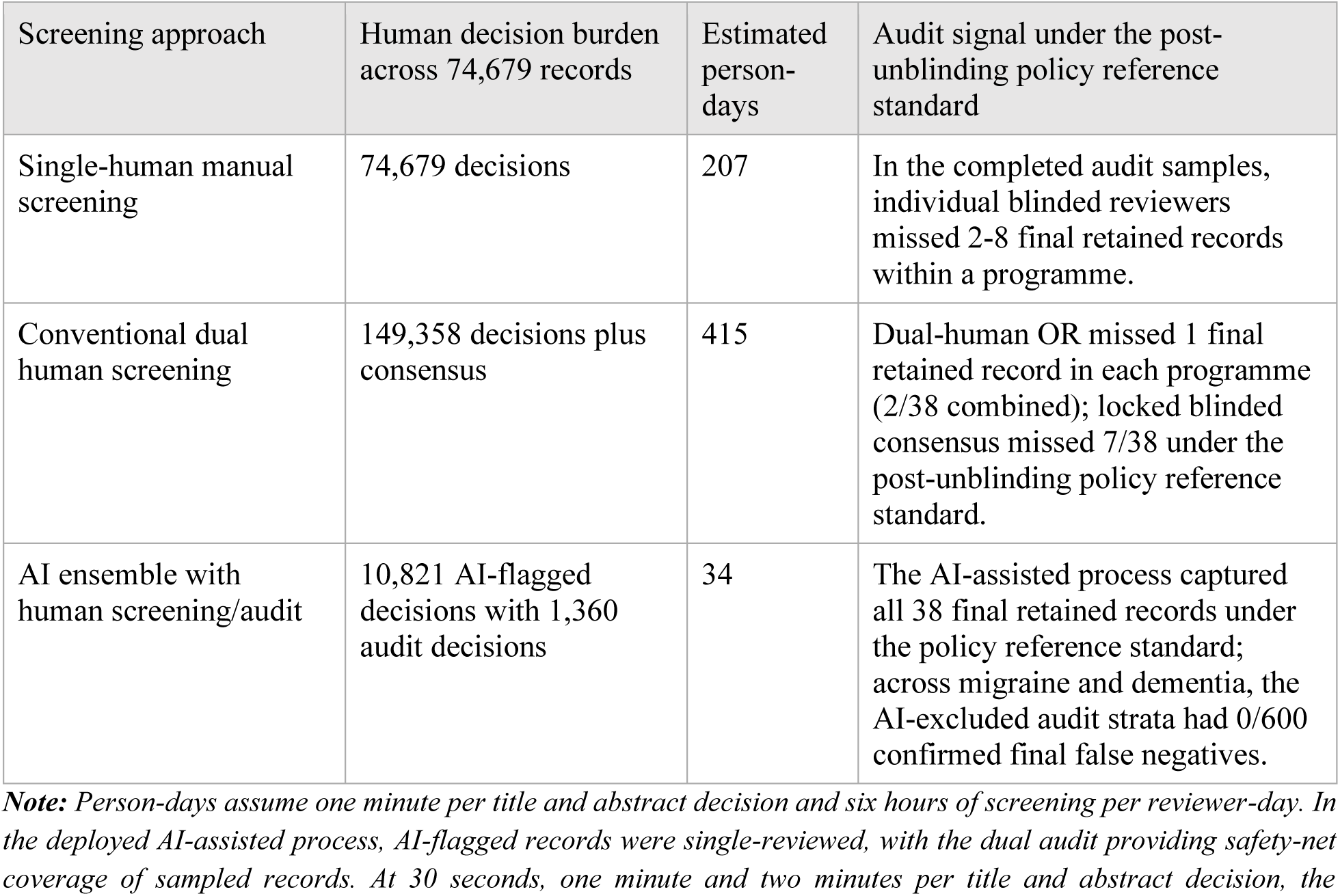
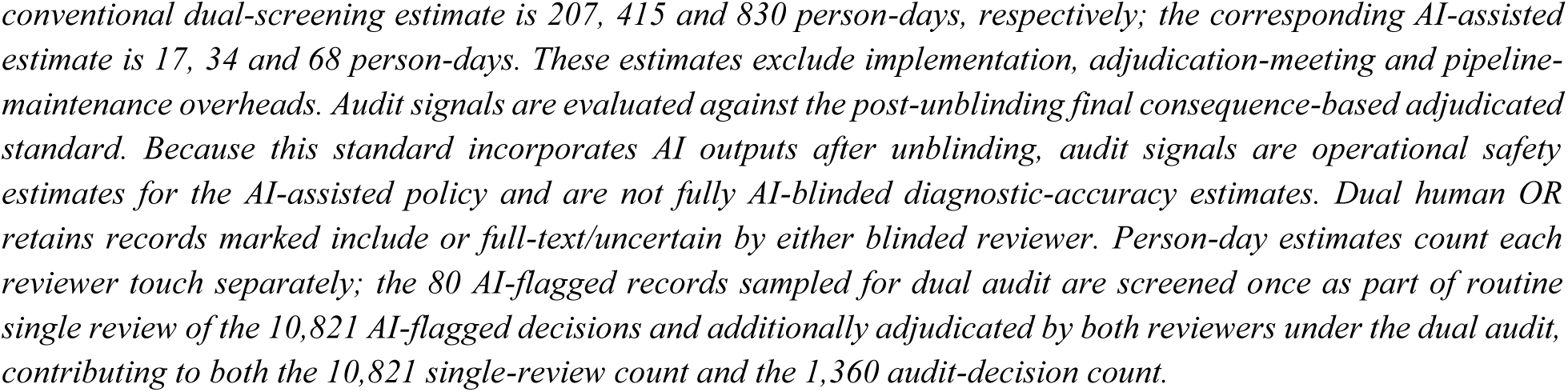
Operational workload and prospective adjudication signals across screening approaches.

| Screening approach | Human decision burden across 74,679 records | Estimated person-days | Audit signal under the post-unblinding policy reference standard |
| --- | --- | --- | --- |
| Single-human manual screening | 74,679 decisions | 207 | In the completed audit samples, individual blinded reviewers missed 2-8 final retained records within a programme. |
| Conventional dual human screening | 149,358 decisions plus consensus | 415 | Dual-human OR missed 1 final retained record in each programme (2/38 combined); locked blinded consensus missed 7/38 under the post-unblinding policy reference standard. |
| AI ensemble with human screening/audit | 10,821 AI-flagged decisions with 1,360 audit decisions | 34 | The AI-assisted process captured all 38 final retained records under the policy reference standard; across migraine and dementia, the AI-excluded audit strata had 0/600 confirmed final false negatives. |

**Note:** Person-days assume one minute per title and abstract decision and six hours of screening per reviewer-day. In the deployed AI-assisted process, AI-flagged records were single-reviewed, with the dual audit providing safety-net coverage of sampled records. At 30 seconds, one minute and two minutes per title and abstract decision, the
*conventional dual-screening estimate is 207, 415 and 830 person-days, respectively; the corresponding AI-assisted estimate is 17, 34 and 68 person-days. These estimates exclude implementation, adjudication-meeting and pipeline-maintenance overheads. Audit signals are evaluated against the post-unblinding final consequence-based adjudicated standard. Because this standard incorporates AI outputs after unblinding, audit signals are operational safety estimates for the AI-assisted policy and are not fully AI-blinded diagnostic-accuracy estimates. Dual human OR retains records marked include or full-text/uncertain by either blinded reviewer. Person-day estimates count each reviewer touch separately; the 80 AI-flagged records sampled for dual audit are screened once as part of routine single review of the 10,821 AI-flagged decisions and additionally adjudicated by both reviewers under the dual audit, contributing to both the 10,821 single-review count and the 1,360 audit-decision count.*

### Prospective adjudication in migraine and dementia

The prespecified 340-record audits were completed for both guideline programmes against the original eligibility criteria (audit flows shown for migraine in Supplementary Fig. 3 and dementia in Supplementary Fig. 4). In each programme, two independent methodologists screened the audit records while blinded to AI outputs, and locked consensus decisions before AI outputs were unblinded. Inter-rater agreement was 82.6% (Cohen’s kappa = 0.32) for three-category decisions and 86.2% (kappa = 0.44) for binary retain versus exclude in migraine; corresponding dementia values were 90.0% (kappa = 0.23) and 90.0% (kappa = 0.23). These kappas reflect the deliberately enriched, low-prevalence audit samples rather than routine full-corpus screening agreement; under the audit’s approximately 5% positive prevalence, kappa is mechanically suppressed even at high raw agreement (see Supplementary Notes 3-5 for the kappa caveat and programme-specific adjudication detail).

Across the 600 sampled AI-excluded records, 19 records were retained at blinded consensus (17 migraine; 2 dementia), all as cautious full-text assessment rather than include. After AI outputs were unblinded, all 19 were reclassified as ineligible after structured criterion-specific adjudication; none was confirmed as a final false negative or judged likely to change a review’s conclusions. Seven AI-flagged records were reclassified from excluded to retained after unblinding because the AI had flagged records initially excluded by blinded human consensus (2 migraine; 5 dementia). This post-unblinding policy reference standard therefore found 0 of 600 sampled AI-excluded false negatives, while the AI-assisted process flagged all 38 final retained records in the combined audit sample (24 migraine; 14 dementia).

Human-comparator performance contextualised these findings against the post-unblinding final adjudicated policy standard (Table 2). In migraine, individual blinded reviewers missed 2 and 5 of 24 final retained records, locked blinded consensus missed 2, and dual human inclusive OR rule missed 1. In dementia, individual blinded reviewers missed 8 and 2 of 14 final retained records, locked blinded consensus missed 5, and dual human OR missed 1. These comparisons describe the policy-evaluation reference standard, not a fully AI-blinded diagnostic-accuracy standard. The AI-assisted process reached 100.0% sample sensitivity in both programmes because no final retained record came from the AI-excluded stratum and met the prespecified 5-percentage-point non-inferiority margin against locked blinded consensus in both programmes. Inverse probability weighted specificity was 92.0% (bootstrap 95% CI, 89.8-94.5) in migraine, 90.6% (88.7-92.9) in dementia and 91.3% (89.8-93.1) combined.

**Table 2.** Layered screening performance in completed prospective adjudication samples.

| Programme and decision layer | Sample TP | Sample FP | Sample FN | Sample TN | Sample sensitivity | Sample specificity |
| --- | --- | --- | --- | --- | --- | --- |
| **Migraine** |  |  |  |  |  |  |
| Reviewer A, blinded | 22 | 38 | 2 | 278 | 91.7% | 88.0% |
| Reviewer B, blinded | 19 | 18 | 5 | 298 | 79.2% | 94.3% |
| Dual human OR, blinded | 23 | 49 | 1 | 267 | 95.8% | 84.5% |
| Locked blinded consensus | 22 | 17 | 2 | 299 | 91.7% | 94.6% |
| AI-assisted process | 24 | 16 | 0 | 300 | 100.0% | 94.9% |
| **Dementia** |  |  |  |  |  |  |
| Reviewer C, blinded | 6 | 3 | 8 | 323 | 42.9% | 99.1% |
| Reviewer D, blinded | 12 | 25 | 2 | 301 | 85.7% | 92.3% |
| Dual human OR, blinded | 13 | 27 | 1 | 299 | 92.9% | 91.7% |
| Locked blinded consensus | 9 | 2 | 5 | 324 | 64.3% | 99.4% |
| AI-assisted process | 14 | 26 | 0 | 300 | 100.0% | 92.0% |
| **Combined programmes** |  |  |  |  |  |  |
| AI-assisted process | 38 | 42 | 0 | 600 | 100.0% | 93.5% |

**Note:** Sample counts and sample performance are unweighted and evaluated against the post-unblinding final consequence-based adjudicated standard. Dual human OR rule retains a record if either blinded reviewer marked it as include or full-text before consensus. The final AI-assisted process had 100.0% inverse-probability-weighted sensitivity in migraine, dementia and the combined audit; inverse-probability-weighted specificity was 92.0% (89.8-94.5), 90.6% (88.7-92.9) and 91.3% (89.8-93.1), respectively. TP, true positive; FP, false positive; FN, false negative; TN, true negative.

## Discussion

To our knowledge, this is the first study to report an LLM screening process selected, validated and prospectively deployed under explicit audit and version control within a national guideline-producing agency. The selected ensemble removed most records from first-pass title and abstract screening while retaining high validation sensitivity. In the completed prospective audits, no sampled AI-excluded record in either migraine or dementia was confirmed as a final false negative after consequence-based adjudication; in both programmes, the AI-assisted process also retained records initially excluded by blinded human reviewers.

Prior work has established that LLM-based screening can be sensitive, that prompt design is important, and that workload reduction can be substantial^15–21,24^. Our earlier studies identified batch-100 inference and dual-model dual-run ensembling as practical requirements for deployment^16,20^. The present study adds three elements: a frozen model pair and prompt carried unchanged from selection through deployment, a workload effect estimated on real search records rather than curated benchmark labels, and prospective guideline use coupled to prespecified audit. Study within a review designs^25^ and adaptive-platform protocols^8^ treat LLM automation as an intervention used inside a single review or across a structured platform; the Cochrane CESAR protocol^8^ in particular sets out a staged plan to evaluate AI for title and abstract screening, full-text screening, and data extraction. This study contributes prospective deployment evidence for the first of those three steps within a national-guideline programme. The 419-review retrospective benchmark also represents a large single-task LLM validation in evidence synthesis. It was evaluated against labels derived from Cochrane reviews. These labels draw on hundreds of reviewer-years of systematic review work across the full spectrum of Cochrane review groups^1,11^.

Because the screening system makes exclusion decisions traceable in a way routine large-scale manual screening typically does not, it also creates two opposing risks: automation bias, in which reviewers defer too readily to AI, and algorithm aversion, in which reviewers dismiss AI outputs despite relevant signals. For example, automation bias has been documented in AI-assisted clinical decision-making, including a recent randomised trial in physicians trained in AI literacy^26^. The audit was designed to minimise both. Each 340-record audit pool per guideline mixed 300 AI-excluded with 40 AI-flagged records, so blinded human reviewers could not infer the AI’s decision from pool membership, and reviewers locked their decisions before AI outputs were unblinded. Reclassification of a blinded-lock positive then required adjudicators to identify a criterion-specific mismatch rather than defer to the AI, and human decisions were themselves evaluated against the final adjudicated policy standard. At deployment scale, this provides a middle ground between full manual rescreening and unvalidated automation: a bounded audit cost, explicit statistical assurance and retained human accountability^9^.

The completed prospective adjudication illustrates why the audit layer matters. Across both programmes, 19 AI-excluded records were retained at blinded consensus but reclassified after AI outputs were unblinded; none remained finally retained or likely to change a review’s conclusions. Conversely, seven AI-flagged records initially excluded by blinded human consensus were retained after criterion-specific adjudication. This bidirectional pattern is important: the AI layer did not merely replace human screening, but created a structured second look at the decisions most likely to matter.

At the same time, the final audit endpoint is not independent of the AI system: final classification was determined after AI decisions and reasons were unblinded. We therefore interpret the prospective audit as an operational evaluation of an AI-assisted screening and adjudication policy, not as a fully AI-blinded diagnostic-accuracy study. The locked blinded consensus metrics, the dual-human OR comparator and the post-unblinding final adjudication are reported separately for this reason. External blinded adjudication of reclassified cases, or complete duplicate human screening of all AI-excluded records, would further strengthen causal claims about AI versus human accuracy.

The retrospective benchmark counts of 82 missed final-inclusion records on 419-SR and 25 in topic-matched external validation should be interpreted in light of Cochrane’s label structure. Cochrane final-inclusion sets may include companion reports, protocols, secondary analyses, and related records for the same underlying study; conversely, published review decisions may themselves reflect human screening omissions. In our earlier 16-review benchmark, a pre-specified adjudication of LLM-Cochrane disagreements found that records where both models disagreed with a Cochrane ’include’ label were in many cases, on re-review against the original eligibility criteria, probable misclassifications in the Cochrane EndNote libraries rather than LLM errors16,20. We did not repeat that adjudication for retrospective AI-label discordances here in the larger 419-SR dataset, because the study aim was to validate the deployed ensemble rather than to recompute Cochrane’s inclusion decisions, so the proportion that represent companion records, consequential primary-study losses, or arguable reference-standard errors is not quantified at this scale. The prospective audits therefore provide the more policy-relevant safety evidence for the deployed process; the same approach could in principle be applied to retrospective error correction in published reviews.

Three findings are particularly relevant for future implementation. First, the combined audit found no confirmed final false negatives in 600 AI-excluded records, but this is an audited upper-bound statement, not proof that misses cannot occur. Second, no-abstract records and unclear guideline, recommendation, consensus or health-technology-assessment records remain safety-relevant because their evidence basis is difficult to assess from title alone; these should be routed directly to human review when systematic reviews or trials are eligible. Third, the specificity cost of conservative routing is visible in the AI-flagged stratum: the system deliberately routed many finally excluded records to human reviewers to protect sensitivity.

The ethical comparison here is not autonomous AI versus an idealised human gold standard but a comparison between alternative screening policies under resource constraints. Full dual human screening maximises redundancy but carries substantial opportunity cost; single human screening is less costly than dual human screening but may miss records that another reviewer or the AI-assisted audit would retain. Delayed guideline updates are themselves a clinical harm. Contemporary systematic reviews already depend on semi-automated infrastructure: database indexing, methodological filters, deduplication algorithms, screening platforms, and human screening under fatigue and time pressure. The distinctive contribution of the present approach is that it makes a high-volume evidence-selection step explicit, measurable, version-locked, and auditable. The relevant implementation question is whether this policy yields lower consequential error, better transparency, and acceptable workload trade-offs against the realistic alternatives already used in evidence production, while recognising that local validation remains necessary before transfer to new agencies, languages, review types or database ecosystems.

The comparison with BioBERT is conceptually important. The BioBERT values shown here were derived from Chan et al.’s published comparator analyses and were not generated by running BioBERT in the present study. Chan et al.’s supervised comparator on 419-SR and 22-SR was trained on a very large in-domain corpus of labelled Cochrane screening decisions, whereas the present LLM triage approach applies review-specific eligibility criteria directly, without task-specific training data, and returns record-level reasons that can be audited^11,12^. For example, their BioBERT model had high false-negative rates on diagnostic test accuracy review topic under-represented in its training corpus, leaving full generalisation to real-world guideline topics uncertain. We did not observe the same high false-negative pattern with the ensemble approach in the corresponding review topic. This observation may lower the barrier to deployment for organisations without large historical screening corpora on their intended review topics, and may improve portability across review types, provided each deployment topic is audited under the same governance model.

The open-weight ensemble did not fully match the selected commercial ensemble’s sensitivity. The transfer of the same ensembling logic suggests that local models may be useful where data governance, cost, service continuity or local-deployment constraints dominate, but they still require local validation and audit.

The policy stakes are growing. As of mid 2026, commercial AI tools for systematic review screening, evidence extraction and synthesis are increasingly marketed to guideline producers and methodologists, often without standardised retrospective validation or prospective audit. In the current study, sensitivity performance was clearly dependent on validation-driven model selection and ensembling (Extended Data Figure 4). Furthermore, the broader risk of unaudited AI-generated scholarly content is illustrated by a recent study that identified fabricated citations across biomedical literature at scale^27^. The principles are not specific to the models tested. Any AI tool used in evidence production should be evaluated on a broad retrospective benchmark, externally validated on records matched to the deployment topics, deployed under locked versions, and audited in the only stratum that removes human review. Tools that cannot be evaluated this way should not displace human screening in systematic review screening during guideline production.

Environmental cost is a real implementation consideration for guideline producers deploying AI screening at scale. We estimated the present system’s footprint rather than measuring it directly. Using published Gemini Apps production measurements^28^, the four-run, batch-100 system gives a lower floor estimate of ∼1 Wh and ∼1 ml of water per 100 records and a conservative token-scaled upper bound of ∼73 Wh and ∼79 ml per 100 records screened. Dual independent human screening on two enterprise workstations, including laptops and external monitors, would consume approximately 167 Wh per 100 records before office overheads and embodied hardware are considered. These provider-derived estimates may not generalise to scientific batch inference, but they suggest environmental accounting should sit alongside accuracy, workload and governance when AI tools are evaluated for evidence production, consistent with recent GRADE guidance^29^. Direct measurement of cloud and locally deployed open-weight approaches should be a priority for future studies.

Commercial-API operating cost for the prospective deployment was modest by guideline-production standards. At the per-1,000-record cost measured for the selected ensemble in development (Supplementary Table 12), the 74,679-record prospective corpus corresponded to approximately USD 400 in inference spend across both guideline programmes. This should be contrasted with the estimated 207-830 person-day range for conventional dual human screening under 30-second to two-minute per-record assumptions, although those estimates do not include all implementation, audit-meeting or maintenance costs of the AI-assisted workflow.

This study has several limitations. Because the ensemble was selected on the 419-SR benchmark, the 98.0% estimate is conditional on model selection; the topic-matched external validation and the prospective audit provide the independent checks. Ensemble selection was also conditional on the granularity of eligibility criteria supplied to the models. The primary ensemble was selected on 419-SR using broad review-level criteria obtained from Cochrane review abstracts rather than full methods-section criteria. This was a conservative choice for heterogeneous guideline deployment because 419-SR is a large dataset spanning the full Cochrane corpus, whereas 16-SR contains only 16 reviews, and because future reviews may not always have detailed criteria fully operationalised at the point of first-pass triage. In the 16-SR detailed-criteria sensitivity analysis, newer frontier ensembles, including GPT-5.5 plus Gemini-3.1-Pro, maintained sensitivity while improving specificity, while having reduced sensitivity with broad criteria. This suggests that specificity may be recoverable when detailed eligibility criteria and newer models are available, but also that the optimal model pair may depend on criteria specification, dataset size and deployment setting.

The 22-SR real-search workload benchmark used a closely related Gemini-3-flash plus GPT-5 ensemble rather than the final GPT-5.1 deployment pair because that run was completed before GPT-5.1 release; the final selected pair should be rerun on comparable real-search corpora when feasible. Prospective evidence comes from one national agency and two guideline programmes; both audit samples are complete, but the estimates still arise from stratified sampling rather than complete duplicate human screening of every AI-excluded record, and may not generalise without local validation to other agencies, languages, database-access environments, review types or clinical domains. The final prospective adjudication standard was determined after AI outputs were unblinded, so it is appropriate for evaluating the implemented AI-assisted policy but not fully independent for estimating AI versus human diagnostic accuracy; external blinded adjudication of reclassified cases by adjudicators not exposed to the AI rationale, or complete duplicate human screening of every AI-excluded record (which would also produce a fully blinded reference for the AI-excluded stratum), would strengthen causal claims about AI versus human accuracy. Training-data contamination remains possible in any LLM benchmark based on published systematic reviews, although the temporally independent development data, non-monotonic release series behaviour, topic-matched external validation, and prospective deployment on records retrieved after model training cutoffs make simple memorisation an incomplete explanation. The prospective safety result should be interpreted as an audited deployment finding and exact upper-bound statement, not proof that no false-negative AI exclusion occurred in the full corpus.

The system evaluated here was limited to title and abstract screening triage. Full-text screening, risk of bias assessment and structured data extraction require full documents, longer context and different adjudication standards; paywalls, copyright, and automated retrieval at scale may create additional implementation challenges. Any downstream use in later stages of the systematic review process should be treated as a new validation problem with its own prompt lock, version control, auditable outputs, escalation rules and empirical audit of consequential errors^9^. The environmental footprint was estimated from published provider measurements and representative token counts rather than measured directly for the deployed system.

Two further limitations bear on implementation. Formal human-factors and usability evaluation, covering user trust, cognitive workload, training burden and implementation behaviour, was outside the scope of this study and should accompany wider implementation. In addition, because the selected ensemble used proprietary models, their internal training data and decision processes were not inspectable; we therefore relied on locked inputs and outputs, cached decisions, prespecified routing rules, external validation, prospective audit and revalidation triggers rather than on claims of model interpretability.

Additionally, recent commentary has rightly cautioned that AI should not replace expert judgement in evidence synthesis^30^. The system evaluated here was restricted to conservative first-pass routing under a locked policy, with human methodologists retaining responsibility for criteria, audit, adjudication and interpretation. Within that role, the prospective audit suggests that responsible oversight need not always mean full duplication of every routed record; it can also mean disciplined delegation, with narrow scope, fail-safe routing, transparent monitoring and human accountability for the decisions that require judgement.

In conclusion, this staged validation and prospective audit supports further evaluated use of locked and audited LLM-ensemble triage with human oversight to reduce title and abstract screening burden in systematic reviews for national guidelines. Prospective deployment requires frozen prompts and model versions, audit sampling, human fallback, conservative routing of no-abstract and ambiguous records to human screening, and local programme-specific monitoring before transfer to new settings. The evidence here spans retrospective validation, topic-matched external validation, real-search workload estimation with a closely related model pair and completed prospective adjudication across two national guideline programmes.

## Methods

### Study design

This was a staged validation to deployment study linking retrospective validation, ensemble selection, topic-matched external validation, real-search workload estimation, and prospective guideline deployment. The primary retrospective endpoint was review-level sensitivity for final inclusion records (label 1.0) in 419-SR, summarised as the arithmetic mean across reviews with at least one final inclusion. The 419-SR benchmark was used as the primary selection substrate because it is the largest corpus-wide benchmark available to this study, spans 419 Cochrane reviews across 53 Review Groups, and gives substantially broader topical coverage than the temporally independent but small 16-SR development set. This endpoint was chosen because the most consequential operational harm is excluding a study that would ultimately enter the evidence synthesis, and because between-reviewer variation in title and abstract caution dominates the label 0.5 stratum, making records that pass title and abstract screening but are later excluded at full text a noisy validation target. Pooled record-level counts of missed inclusions are reported alongside as operational summaries. The primary prospective endpoint was the false negative proportion in the AI-excluded stratum: records receiving E from all four runs and therefore not sent to first-pass human screening. Reporting was informed by STARD-AI^31^, MI-CLAIM-GEN^32^, and RAISE 2/3 principles^9^ (Supplementary Tables 2-4); under RAISE 3 the system was treated as a generative LLM support tool for study selection requiring validation within the intended review context. National guideline production at the Swedish National Board of Health and Welfare follows agency methodology^23^, complemented by Swedish Agency for Health Technology Assessment and Assessment of Social Services (SBU)^33^ and Cochrane^4^ guidance for systematic review and evidence-grading steps.

The selection design specifically prioritised cross-family ensembling because the most consequential operational harm, missing a study that would ultimately enter the evidence synthesis, is sensitive to model-family bias. Single-family ensembling preserves shared failure modes, including training-data overlap, instruction-tuning conventions, and decoding tendencies. Cross-family ensembling was intended to reduce correlated misses by combining models with different training corpora and post-training procedures. The four-run OR rule then converts this diversity into sensitivity at the cost of specificity, which is acceptable because all flagged records are routed to human review. This sensitivity-first operating point is consistent with Cochrane guidance that search and early study-selection processes should favour high sensitivity and over-inclusion^4^.

### Datasets

Eight dataset groups were used (Table 3): 16-SR (16 Cochrane reviews, May-June 2025, temporally independent of training cutoffs for the main retrospective models; used for development and criteria-granularity sensitivity analyses); 419-SR from Chan et al.^11,12^ (26,892 records, 419 reviews across the Cochrane corpus; primary retrospective validation and ensemble selection); 22-SR also from Chan et al. (142,504 unfiltered real-search records, 22 reviews; workload estimation); topic-matched external migraine (1,343 records, 27 reviews) and dementia (7,158 records, 106 reviews) Cochrane subsets, drawn from the Chan et al.11 training partition and not used in any of the present study’s other validation analyses; four prespecified normatively contested topic subsets (abortion, autism, vaccines and HIV; 9,765 records, 177 reviews), also derived by keyword matching within non-overlapping Chan et al. training-partition reviews and used only for sensitive-topic/bias-oriented robustness analysis (Supplementary Fig. 1); and prospective migraine (36,503 records, 16 reviews) and dementia (38,176 records, 8 reviews) live deployment corpora.

**Table 3.** Datasets and reference standards.

| Dataset | n records | n reviews | n included / positive | Purpose |
| --- | --- | --- | --- | --- |
| 16-SR | 736 | 16 | 329 | Development and prompt / criteria analyses |
| 419-SR | 26,892 | 419 | 4,545 | Primary retrospective validation |
| 22-SR | 142,504 | 22 | 768 | Real-search workload estimation |
| External migraine | 1,343 | 27 | 293 | Topic-matched external validation |
| External dementia | 7,158 | 106 | 671 | Topic-matched external validation |
| Normatively contested topic subsets | 9,765 | 177 | 1,400 | Prespecified sensitive-topic / bias robustness analysis |
| Prospective migraine | 36,503 | 16 | audit sample adjudicated (24 records retained) | Live deployment |
| Prospective dementia | 38,176 | 8 | audit sample adjudicated (14 records retained) | Live deployment |

**Note:** Counts refer to screening records unless otherwise stated. Prospective datasets report post-deduplication record counts. For 16-SR, 329 is the corrected-label positive count used in all analyses (after an author correction to the label\_corrected reference-standard ordering); the original Cochrane label count is 348 and is preserved for traceability in the reproducibility archive.

For retrospective datasets, published review decisions served as the reference standard. For prospective datasets, blinded dual review with locked consensus before AI outputs were unblinded served as the primary human screening reference, followed by consequence-based adjudication for final classification. Eligibility criteria fed to the screening prompt differed across datasets by design. For 419-SR, The 22-SR real-search corpus, the topic-matched external migraine and dementia validation subsets, and the normatively contested topic subsets, criteria were the review-level metadata fields released by Chan et al.^11,12^ (title, background, objectives and selection criteria for each Cochrane review), the same fields used to train and evaluate the BioBERT baseline. The more granular inclusion criteria from each review’s full methods section were not extracted for these datasets because harmonising methods-section criteria across 419 reviews, 22-SR, external-validation subsets and sensitive-topic subsets was not feasible. The broad-criteria 419-SR selection design was retained because it provided a large, general Cochrane-wide test of conservative first-pass triage and approximated the variable level of criteria specification that may be available when future guideline reviews begin title and abstract screening. To quantify the effect of criteria richness, the smaller 16-SR development corpus was screened twice: once with the Chan et al. style review-level metadata (16-SR broad) and once with full methods-section criteria (16-SR detailed). The broad versus detailed comparison is reported as a sensitivity lever in Extended Data Fig. 2, Supplementary Data Table 11 and Supplementary Table 12 and modestly favoured detailed criteria, particularly for GPT-family models. The prospective migraine and dementia deployments used full methods section criteria. The prospective deployment also used a concise variant of the locked system prompt that removed Cochrane-specific routing conventions inapplicable to national guideline production. Cochrane reviews retain study protocols, design papers and trial registry entries describing eligible studies as ongoing/awaiting classification, whereas Swedish national guidelines usually exclude study protocols and ongoing studies and operate on completed evidence only; the prospective variant therefore tightened delivery modality and co-intervention clauses to match guideline review practice. Both system prompt variants are reproduced in Supplementary Note 1, and the per-dataset criteria-source and prompt-variant mapping is given in Supplementary Table 16.

### Models, prompts, and decision rules

Ten LLMs across five model families (Gemini, OpenAI GPT, GPT-OSS, GLM, and Gemma; see Supplementary Table 1) spanning commercial API systems and open-weight systems were evaluated on 419-SR; additional 16-SR criteria-richness and cost-comparator analyses evaluated during development are reported in Supplementary Data Table 11 and Supplementary Table 12. All runs used a single locked screening prompt (Supplementary Note 1) with a three-way include / full-text assessment / exclude (I/F/E) output schema (Table 4) and one-sentence justification. Two independent runs per model were obtained. The operational OR-ensemble rule routed a record to human review if any of the four runs returned I or F; only records receiving unanimous E across all four runs entered the AI-excluded pool. For visualisation and threshold analyses, runs were scored as I = 1.0, F = 0.5 and E = 0.0, then averaged across four runs into percentage buckets in 12.5-pp increments. Operationally, any score above 0% routed the record to human review; only records with 0% entered the AI-excluded pool. Batch size was 100 records per prompt for commercial Gemini and GPT models^16^ and 1 for open-weight models where larger batches degraded schema compliance. The evaluated system was a zero-shot inference pipeline with locked prompts and review-specific eligibility criteria; no retrieval augmentation, fine-tuning, or model-weight updating was used. A model card summarising intended use, inputs, outputs, oversight and reassessment triggers is provided in Supplementary Table 15.

**Table 4.** Core components of audited national-guideline LLM screening.

| Component | Implementation in this study | Why it mattered |
| --- | --- | --- |
| Staged validation | 419-SR benchmark, topic-matched external validation, 22-SR real-search workload benchmark | Prevented direct leap from model promise to live deployment |
| Locked configuration | Frozen prompt, cross-family model pair, batch size, output schema and routing rule | Made the tool auditable and prevented silent drift during deployment |
| Conservative exclusion rule | Records routed to the AI-excluded pool only if all four runs returned Exclude (E) | Prioritised sensitivity over specificity |
| Human review of all AI-flagged records | Any Include (I) or full-text assessment (F) signal from any run routed to human screening | Preserved human responsibility for potentially relevant records |
| Prespecified audit | 300 AI-excluded and 40 AI-flagged records per guideline programme | Checked the stratum not sent to first-pass human review |
| Blinded human consensus before AI outputs were unblinded | Reviewers locked decisions before seeing AI outputs or reasons | Reduced anchoring and automation bias |
| Consequence-based adjudication | Errors graded by whether they were likely to change a review's conclusions | Focused safety assessment on clinically meaningful evidence loss |
| Escalation and fallback | Major errors, repeated patterns, technical failure or model change triggered containment/revalidation | Allowed containment and revalidation |

**Note:** Each record received a decision of Include (I), Full-text assessment (F), or Exclude (E) from each of four LLM runs (two models × two independent runs per model). The any-positive routing rule sent a record to human screening if any of the four runs returned I or F; records were routed to the AI-excluded pool only if all four runs returned E. “Likely to change a review’s conclusions” denotes a missed record that would have altered the review’s conclusion or recommendation under consequence-based adjudication. Components were prespecified before each national-guideline programme.

The deployment ensemble Gemini-3-flash plus GPT-5.1 was selected for the highest validation sensitivity meeting prespecified specificity criteria on the large 419-SR broad-criteria benchmark; release-pair drift on 419-SR is reported in Supplementary Table 13. In the smaller 16-SR detailed-criteria sensitivity analysis, GPT-5.5 plus Gemini-3.1-Pro reached 100.0% sensitivity (0 of 329 retained records missed) versus 99.7% for the selected pair (1 missed), with higher specificity, but this was treated as an exploratory specificity-recovery finding rather than a replacement selection rule because 16-SR contains only 16 reviews and was used as a development dataset rather than as the corpus-wide primary benchmark (Supplementary Table 12). The 22-SR workload benchmark used a closely related dual-family workload-estimation ensemble with GPT-5 in place of GPT-5.1 because the run completed before GPT-5.1 release; release-pair analysis (Extended Data Fig. 5) showed the ensemble-level sensitivity gap was 0.4 pp, but the 22-SR result should nevertheless be interpreted as workload evidence from a near-predecessor pair rather than an exact rerun of the final deployment ensemble.

### Search automation and prospective screening

The prospective programme covered 24 reviews across six guideline question categories in the Swedish national guidelines for migraine and dementia. National guidelines at the Swedish National Board of Health and Welfare are statutory recommendations issued to support clinical and managerial priority-setting across the health system^23^. Searches were executed programmatically using PubMed Entrez E-utilities and the EBSCO EDS API against database-specific strategies in a master search workbook, including CINAHL, PsycINFO, PsycArticles, Cochrane CDSR, and CENTRAL where enabled. Records were assigned source-specific identifiers and deduplicated using rule-based exact matching on normalised DOI, title, year, and source-specific identifiers (full rules in Supplementary Methods; pipeline overview in Supplementary Fig. 2). The process retained human review for all AI-flagged records.

### Prospective adjudication and operational safeguards

Each guideline used a prespecified mixed-stratified audit package of 340 records (300 from the AI-excluded stratum, 40 from the AI-flagged stratum), with AI-excluded records sampled using a review-balanced design that approximated proportional allocation while enforcing a minimum contribution per review. Programme-level inverse-probability estimates used the corresponding within-programme stratum sampling fractions. Each guideline programme was adjudicated by an independent pair of experienced methodologists at the Swedish National Board of Health and Welfare; four methodologists in total across the two programmes, with no overlap between the migraine and dementia audit pairs. All four held doctoral-level qualifications and had several years of systematic review screening experience in guideline production. Within each guideline, the two adjudicators independently screened each sampled record against the original eligibility criteria, blinded to each other and to AI outputs. Reviewers met for consensus, which was locked before AI decisions and reasons were unblinded. To reduce anchoring, reclassification of a record retained at blinded consensus required both adjudicators independently to agree that the AI reasoning identified a specific eligibility-criterion mismatch not captured in the original blinded rationale; records were not reclassified solely because the AI had excluded them. Human methodologists retained responsibility for all final decisions; the AI system was used only as triage and audit support.

The audit used stepped monitoring rules. Adjudicated errors were graded as major (false AI exclusions likely to alter the evidence table, synthesis, certainty, or recommendation), moderate (false exclusions unlikely to alter conclusions), workload errors, including false-positive retains and routing decisions, and inconsequential (rationale disagreements). A pooled major AI exclusion error proportion exceeding 3%, or a lower 95% exact CI bound exceeding 2%, was prespecified to trigger escalation to senior methodological review of the affected programme, drawing on the CESAR adaptive platform protocol^8^ as a reference for stepped harm monitoring rather than as an automatic stop rule. Programme-level response, including possible suspension, was reserved for senior methodologists. A 5-percentage-point sensitivity margin against locked human consensus, prespecified to reflect the clinically meaningful loss of sensitivity below which AI-assisted screening would not be acceptable, was used for non-inferiority monitoring. The AI-assisted policy was compared with each blinded reviewer and with dual human OR and locked consensus, using single human screening as the primary operational comparator. Heightened oversight was triggered by any audited AI-excluded record adjudicated positive. The suggested audit monitoring and stepped escalation rules are provided in Supplementary Table 14.

### Prospective endpoint definition and adjudication sequence

The prospective audit endpoint was defined as the false-negative proportion in the AI-excluded stratum under the post-unblinding final adjudicated standard, with the corresponding Clopper-Pearson upper bound. The adjudication sequence was: (1) blinded dual reviewers screened each audit record against the original review eligibility criteria, blinded to each other and to AI outputs; (2) the two adjudicators met for consensus and the consensus decision was locked to disk before AI outputs were unblinded; (3) AI decisions and reasons were then unblinded; (4) records where AI disagreed with locked consensus were re-screened against the original eligibility criteria, and reclassification of a record retained at blinded consensus required both adjudicators independently to agree on a criterion-specific mismatch not captured in the original blinded rationale, with records not reclassified solely because the AI had excluded them; (5) the resulting final adjudicated decisions are the reference standard for the prospective endpoint reported in this manuscript. Because step (5) incorporates information from the AI system, this endpoint evaluates the AI-assisted screening and adjudication policy as a whole rather than estimating AI versus human diagnostic accuracy independently.

### Outcomes, comparator handling, and statistical analysis

For performance generalisation, the primary estimand was the systematic-review-level mean of the metric of interest. Sensitivity and specificity were first calculated within each review, and the reported point estimate is the arithmetic mean across reviews with at least one record contributing to the denominator for that metric. Uncertainty was quantified using nonparametric bootstrap resampling of reviews (B = 2,000), recalculating the review-level mean in each replicate and taking the 2.5th and 97.5th percentiles; for each estimand the bootstrap resampled only the eligible reviews: reviews with at least one included record for sensitivity and reviews with at least one non-included record for specificity, so that the effective denominator was constant across replicates. For 419-SR this corresponds to 361 sensitivity-eligible reviews of 419 and 419 specificity-eligible reviews; for the combined external validation, 112 sensitivity-eligible reviews of 133. Unless explicitly labelled as pooled record-level (micro) values, all 419-SR model-comparison sensitivity and specificity percentages, including the open-weight comparators, use this same review-level (macro) estimand. The systematic review, not the individual record, was the bootstrap unit because records within a review share eligibility criteria, topic, search strategy, and screening difficulty. Operational quantities, including records excluded, records routed to human review, workload reduction, and absolute missed included studies, were reported as pooled record-level counts because they describe screening burden rather than cross-review generalisation. Throughout the manuscript, performance percentages with intervals are mean review-level estimates with review-clustered bootstrap percentile intervals; counts and workload statements are pooled. Exact binomial confidence intervals were used only for the prespecified prospective harm-monitoring boundary for major AI-exclusion errors and for the audit upper bound on the AI-excluded false-negative rate. Within-model stability was quantified using Cohen’s kappa between independent runs. Exploratory sensitivity-lever contrasts (provider family, OR-ensembling, criteria richness, cross-family pairing) were prespecified descriptive contrasts with paired review-clustered bootstrap CIs within each factor contrast (Extended Data Fig. 2) and were not corrected for multiplicity. We additionally summarised ensemble-score threshold trade-offs on 419-SR (Extended Data Fig. 8).

For prospective adjudication, sensitivity and specificity were estimated using inverse-probability weighting with programme- and stratum-specific inclusion probabilities. AI-excluded strata used inverse probabilities corresponding to 300/30,921 sampled migraine records and 300/32,937 sampled dementia records. AI-flagged strata used inverse probabilities corresponding to 40/5,582 sampled migraine records and 40/5,239 sampled dementia records, with flagged records stratified by ensemble inclusion-score bucket for specificity estimation. Unweighted sample counts are reported separately from inverse-probability-weighted estimates. Bootstrap intervals for prospective inverse-probability-weighted specificity were obtained by stratified bootstrap (B = 2,000) with replacement within the AI-excluded and AI-flagged audit strata, holding stratum-specific inclusion probabilities fixed. The AI-excluded false-negative safety bound is reported separately as a one-sided exact (Clopper-Pearson) 95% interval for the 0/300 per-programme and 0/600 combined audit results, yielding upper bounds of approximately 1.0% per programme and 0.5% combined. Prospective performance estimates use the post-unblinding final consequence-based adjudicated standard; blinded-lock metrics are reported separately to show the pre-unblinding human screening state. Per-review prospective screening statistics are reported in Supplementary Data Tables 6-7; reviewer and decision-layer accuracy, AI confusion matrices, and post-unblinding revisions are retained as audit-workbook provenance rather than as duplicate supplementary tables.

BioBERT comparator performance was not generated in the present study; it was derived from Chan et al.^11^ for the label-1.0 endpoint (their Table 2: 97.5% record-level sensitivity and 65.0% record-level specificity on 419-SR; 70.9% record-level specificity on 22-SR; review-level estimates were not available because record-level predictions were not released by the comparator publication). Workload estimates assumed one minute per title and abstract decision and six hours of screening per reviewer-day in the base case, with 30-second and two-minute per-record sensitivity estimates reported alongside the prospective workload results.

### Ethics

This study did not involve recruitment of human research participants, patient-level data, identifiable personal data, biological material, clinical images, clinical interventions, or patient-care decisions. The work processed only bibliographic metadata (titles, abstracts, identifiers, and review-level eligibility criteria) from published systematic reviews and operational guideline searches. No patient-level data, identifiable individual data, full-text articles, author names, reviewer identities, or adjudication rationales were submitted to model APIs. The evaluated system was intended only for title and abstract triage within systematic review production. It was not used as a patient-facing clinical tool, diagnostic system, treatment recommendation system, or autonomous guideline-authoring system. Final inclusion decisions and guideline recommendations remained under human responsibility: all potentially relevant records were subject to human review or prespecified audit, and human methodologists retained responsibility for final study-selection decisions. Any change in model version, provider, prompt, batch size, output schema, eligibility criteria class, or deployment domain was treated as a new system version requiring revalidation before routine use.

### LLM assistance

OpenAI Codex and Anthropic Claude Code assisted with code development for the inference pipeline, figure generation, and statistical analysis scripts; all figures were generated programmatically from source-data files using this code, and all outputs were reviewed by the authors. Generative AI was not used to generate scientific claims, design the study, make adjudication decisions, or interpret findings. The authors take responsibility for the final manuscript, code, analyses, and interpretation.

## Acknowledgements

We thank the Chan et al. research group for making the underlying screening dataset publicly available^12^ and the Cochrane Collaboration for maintaining the evidence base used in the evaluation.

## Funding

This work was conducted as part of the authors’ employment at the Swedish National Board of Health and Welfare; no external grant funding was received.

## Author contributions

P.F.: conceptualisation, methodology, software, formal analysis, investigation, data curation, writing original draft, visualisation. O.S., K.V.P., C.T. and L.L.: methodology, investigation, validation, writing review and editing. A.B., A.N., N.B. and T.L.: supervision, writing, review and editing. All authors approved the final version of the manuscript.

## Competing interests

The authors declare no financial competing interests related to AI model vendors, evidence-synthesis software vendors, or bibliographic database providers. Authors are employed by the Swedish National Board of Health and Welfare (Socialstyrelsen), which conducts national clinical guideline evidence synthesis and may benefit operationally from improved screening workflows.

## Data availability

The figure source data and supplementary data tables are provided with this manuscript. The full reproducibility archive, comprising record identifiers, review-level metadata, locked prompts, model identifiers and settings, cached model outputs, reference-standard labels, adjudication decisions, and analysis code, will be deposited in a public Zenodo repository under CC BY 4.0 (data) and the MIT License (code). The 419-SR, 22-SR, and external migraine and dementia validation datasets originate from Chan et al.^11^ and are publicly available under CC BY 4.0 through the Mendeley repository^12^ (https://data.mendeley.com/datasets/7sgmg89zb6/1). Full abstracts retrieved from NCBI E-utilities and from EBSCO Discovery Service against CINAHL, PsycINFO, PsycArticles, Cochrane CDSR and Cochrane CENTRAL for the prospective migraine and dementia programmes are subject to publisher and aggregator licence terms and are not redistributed in the public deposit; record identifiers (DOI and PMID where available) are provided so that abstract text can be retrieved under the reader’s own database subscriptions. No patient-level data, identifiable individual data, full-text articles, author names, or reviewer identities were collected, processed, or shared.

## Code availability

All analysis code, inference scripts, prompt templates, model-version metadata, inference settings, audit templates, smoke tests, and adjudication materials will be deposited in a public Zenodo repository, with the code released under the MIT License and the prompts, templates, and adjudication materials released under CC BY 4.0. The reproducibility archive contains the analysis pipeline needed to regenerate the headline statistics, figure source data, supplementary tables, and figure files from the cached LLM outputs included in the archive. Exact re-execution of LLM inference is not guaranteed because external model backends can change over time; the archive therefore includes the prompts, model identifiers, inference settings, inference dates, and cached responses used for the manuscript analyses. AI tools assisted code drafting and editing; analysis code was reviewed and tested by the authors.

## Extended Data

**Extended Data Fig. 1.**
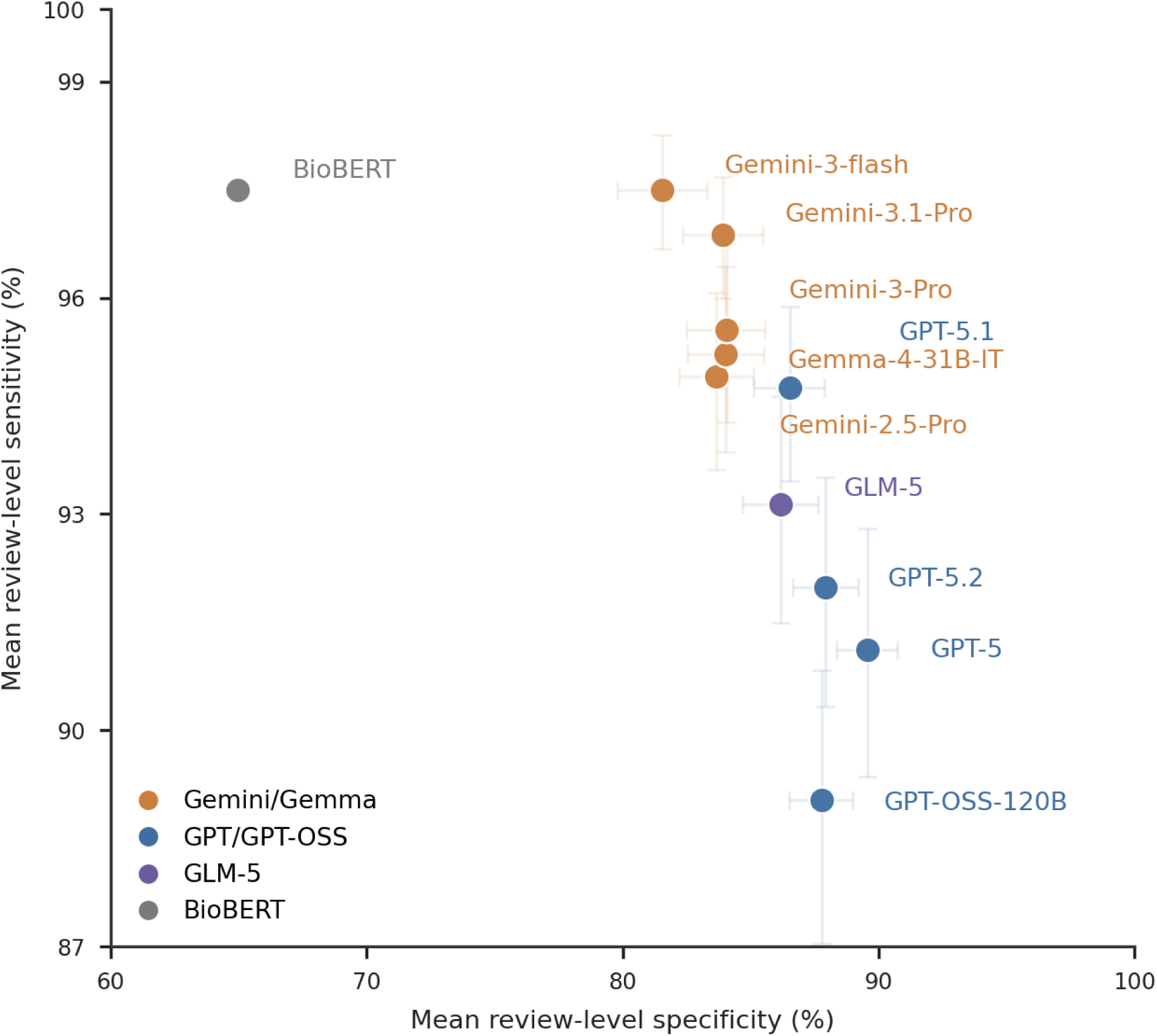
Candidate model performance on the 419-SR validation set. Mean review-level sensitivity versus mean review-level specificity for the ten individual candidate LLMs evaluated on the 419-SR validation set, together with the BioBERT comparator. Points show mean review-level estimates; horizontal and vertical bars show 95% review-clustered percentile bootstrap intervals for specificity and sensitivity, respectively. BioBERT is retained as an external supervised baseline; review-level intervals are not shown for BioBERT because record-level predictions were not available from the comparator publication. Models are coloured by family: Gemini/Gemma, GPT/GPT-OSS, GLM-5, and supervised baseline. This figure provides the detailed candidate-model operating-space view corresponding to the model-selection process summarized in Fig. 1.

**Extended Data Fig. 2.**
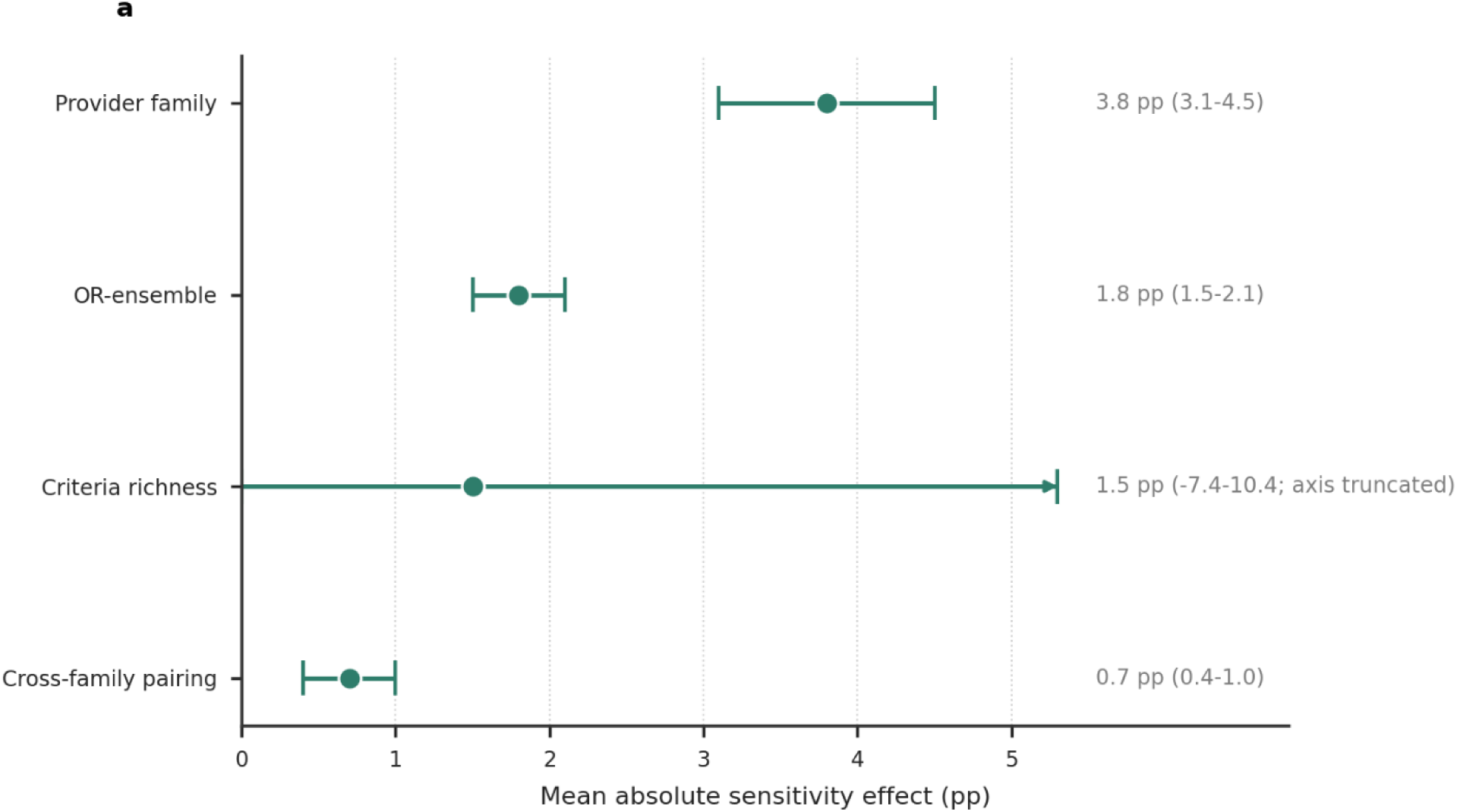
Relative effects of model family, ensembling, and criteria specification. Mean absolute sensitivity effects with paired review-clustered bootstrap 95% intervals. Provider family had the largest average effect (3.8 percentage points; 3.1-4.5), exceeding OR-ensembling (1.8 pp; 1.5-2.1), criteria richness (1.5 pp; -7.4-10.4; axis truncated), and cross-family pairing (0.7 pp; 0.4-1.0). Criteria-richness estimates are based on the 16-SR broad versus detailed comparison; the other sensitivity levers are based on 419-SR.

**Extended Data Fig. 3.**
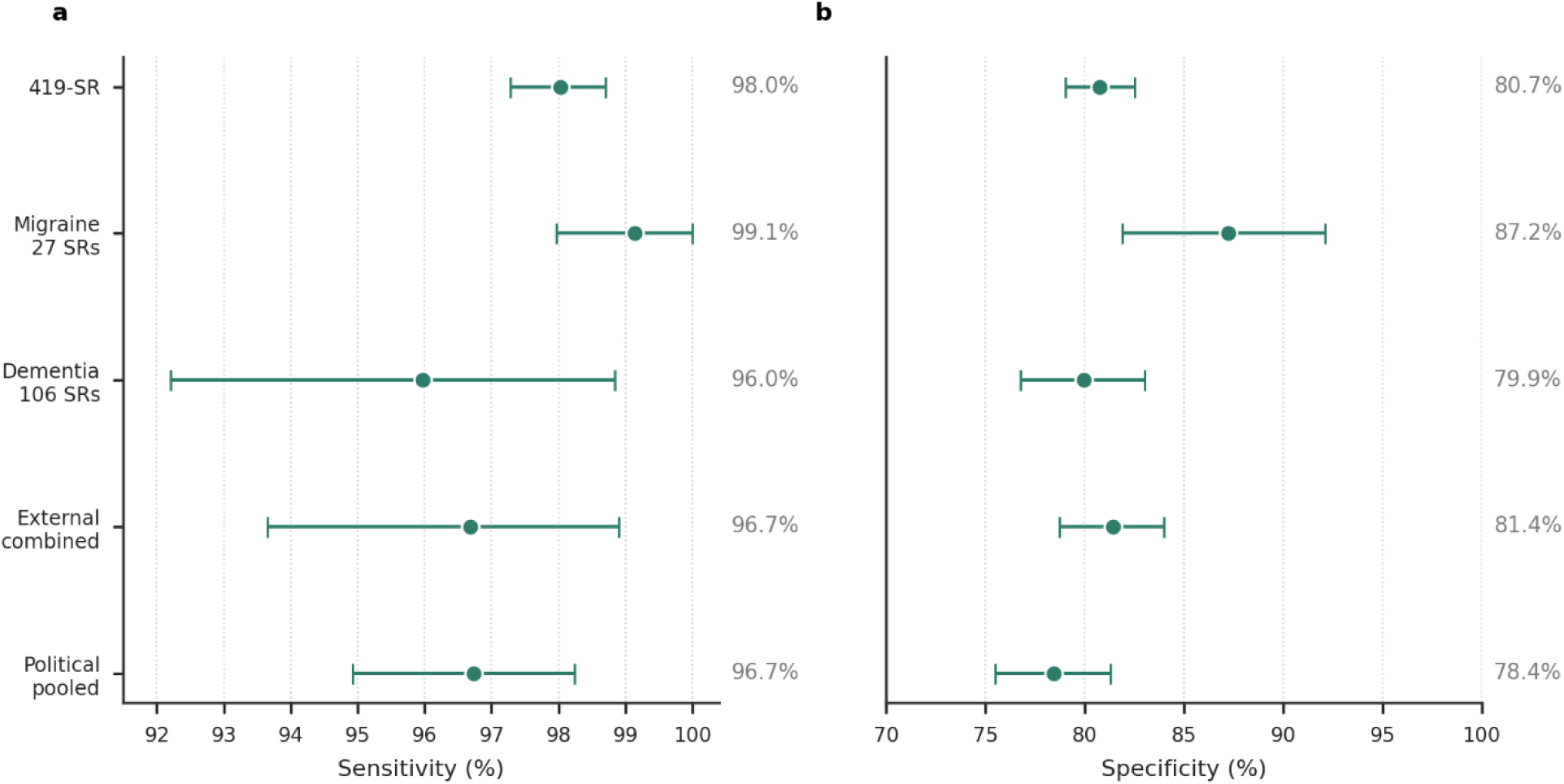
Topic-matched external validation and topic-shift generalisation of the selected LLM ensemble. a, Mean review-level sensitivity of the selected Gemini-3-flash plus GPT-5.1 ensemble across 419-SR, external migraine reviews, external dementia reviews, the combined external migraine and dementia set, and normatively contested topic datasets (abortion, vaccines, autism, and HIV review topics). b, Corresponding mean review-level specificity. Points show review-level means and whiskers show 95% review-clustered bootstrap percentile intervals.

**Extended Data Fig. 4.**
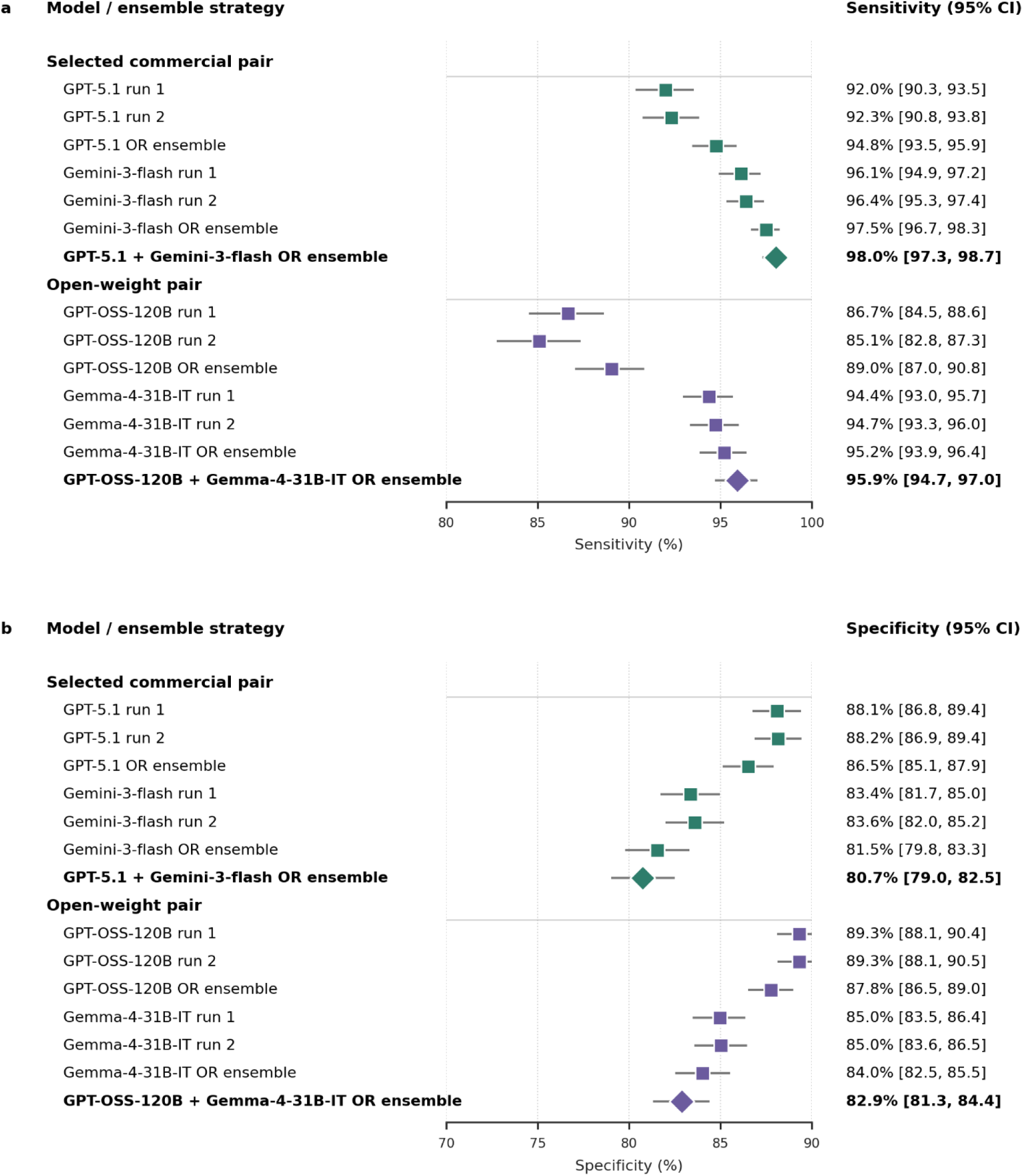
Run-level and ensemble-level performance of selected commercial and open-weight model pairs on 419-SR. a, Sensitivity. b, Specificity. Squares show individual runs and within-model OR ensembles; diamonds show final dual-model, dual-run OR ensembles. Points are mean review-level sensitivity (a) and mean review-level specificity (b); horizontal bars show 95% review-clustered bootstrap percentile intervals.

**Extended Data Fig. 5.**
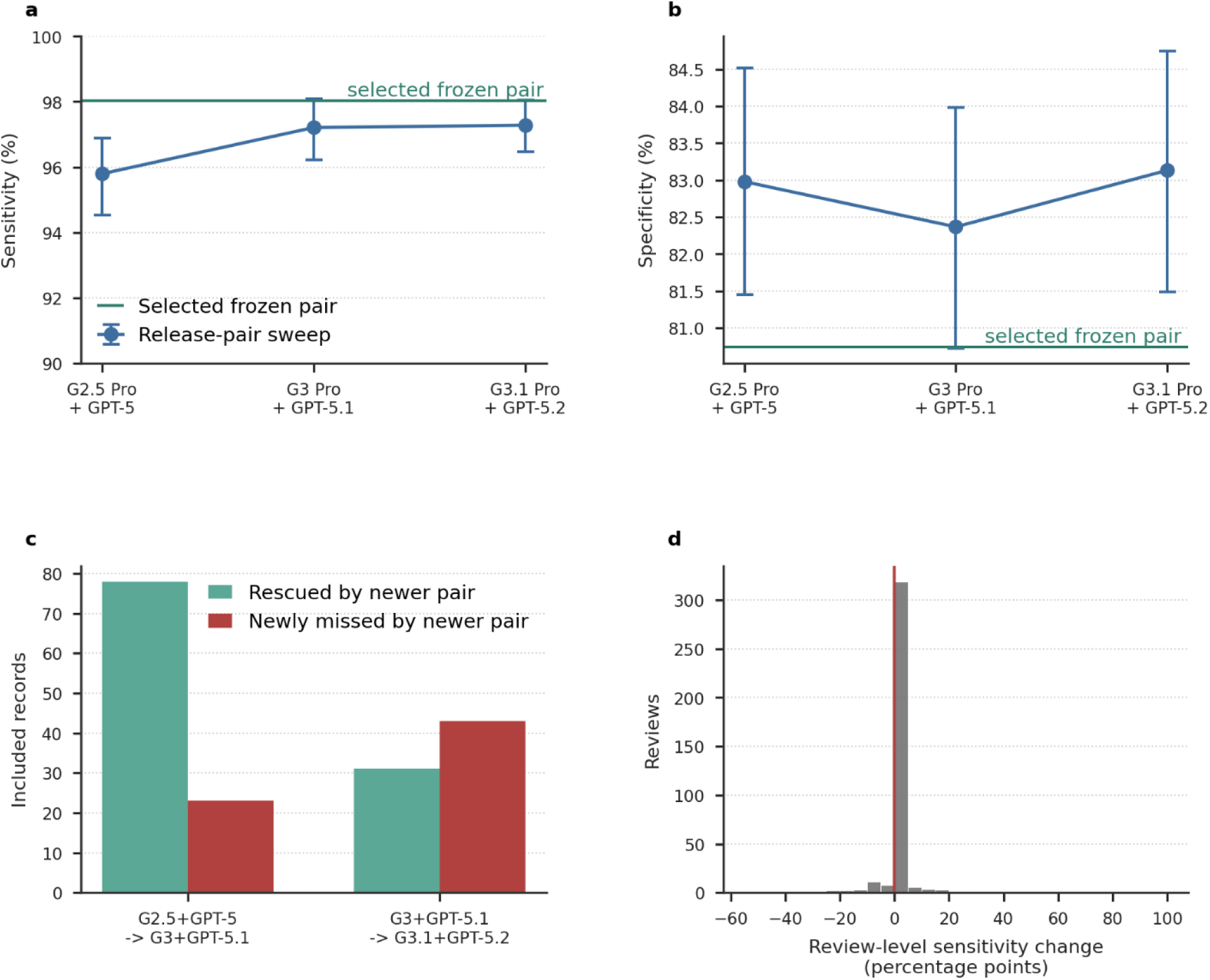
Release-pair drift under dual-model, dual-run ensembling on 419-SR. a, b, Mean review-level sensitivity (a) and specificity (b) for temporally ordered Gemini Pro plus GPT release-pair ensembles on the 419-SR final-inclusion endpoint. Points are review-level means; error bars show 95% review-clustered bootstrap percentile intervals. Horizontal lines show the selected frozen deployment ensemble, Gemini-3-flash plus GPT-5.1, which was not part of the Gemini Pro release-pair trajectory and is therefore shown as a reference rather than connected to the release sequence. c, Included-record migrations between adjacent release-pair ensembles (pooled record-level counts of records rescued and records newly missed by the newer pair). d, Distribution of review-level sensitivity change from Gemini-3-Pro plus GPT-5.1 to Gemini-3.1-Pro plus GPT-5.2 among reviews with at least one included record. The analysis uses the operational unanimous-exclusion rule: records were routed to human review if any of the four runs returned Include or Full-text assessment; only unanimous exclusions were placed in the AI-excluded pool.

**Extended Data Fig. 6.**
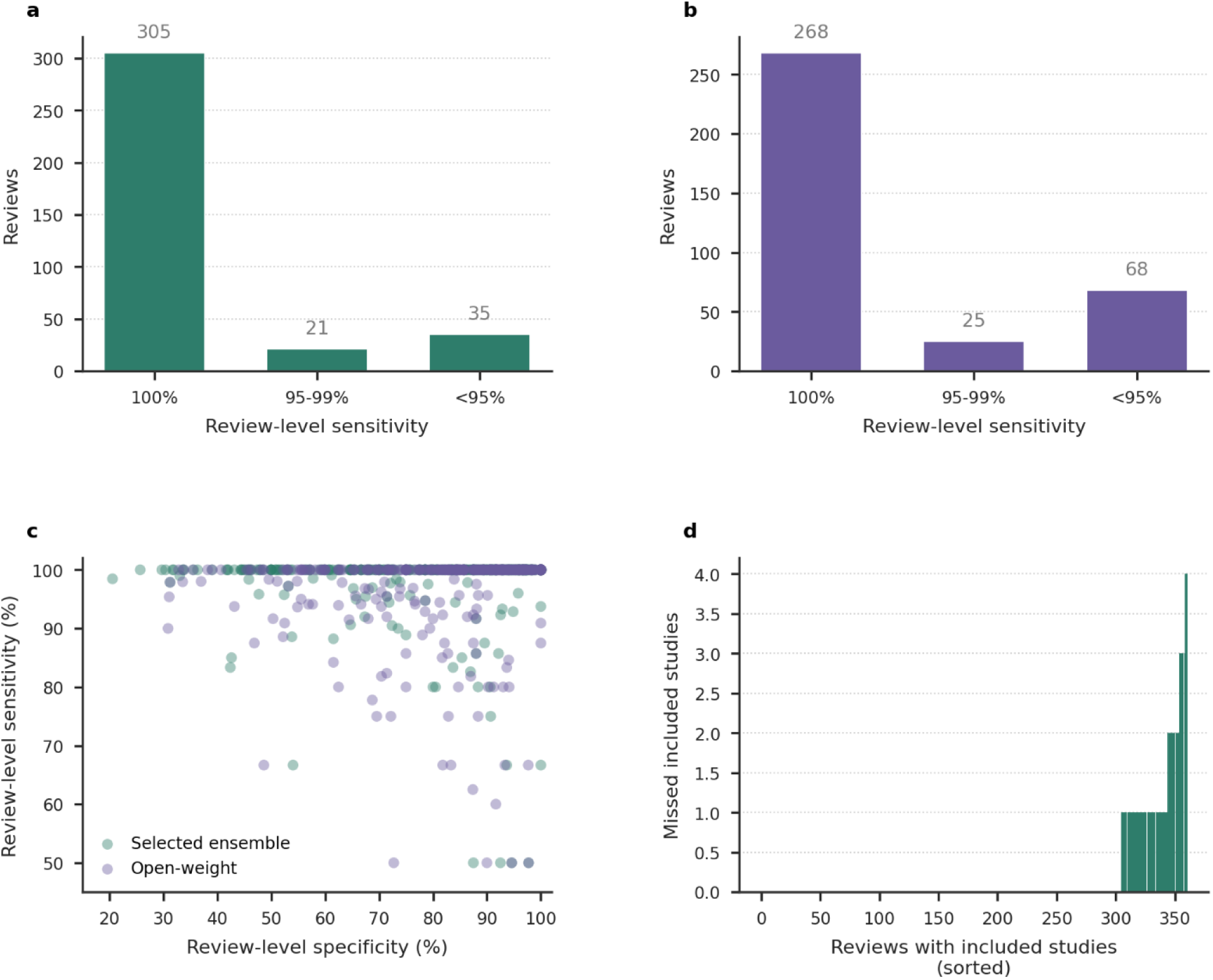
Per-review ensemble performance on 419-SR. a, Reviews with no missed included records for the selected Gemini-3-flash plus GPT-5.1 ensemble. b, Reviews with no missed included records for the open-weight Gemma-4-31B-IT plus GPT-OSS-120B ensemble. c, Review-level sensitivity versus specificity. d, Missed final-inclusion records per review for the selected ensemble. Per-review mean sensitivity (macro-average) summaries corresponding to these distributions are reported in Supplementary Table 10.

**Extended Data Fig. 7.**
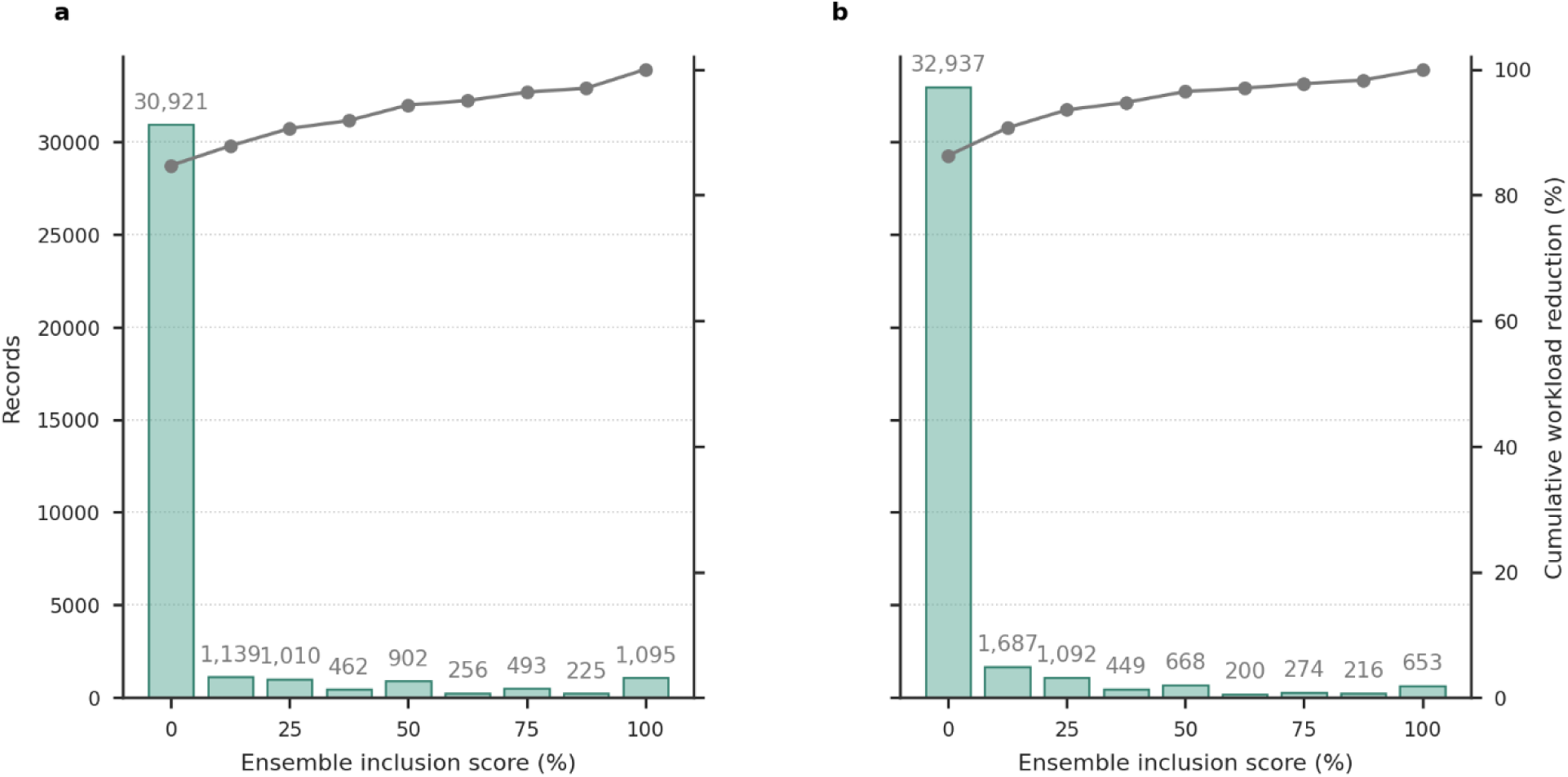
Ensemble inclusion score distributions in prospective migraine and dementia deployments. a, Migraine deployment records (n = 36,503) by four-run ensemble inclusion score bucket. b, Dementia deployment records (n = 38,176) by the same score. The ensemble inclusion score is a deterministic triage score, not a calibrated probability. Each model run was encoded as I = 1.0, F = 0.5, E = 0.0; scores were averaged across four runs and expressed as percentages. A 0% score corresponds to unanimous exclusion (E/E/E/E). A 12.5% score is the lowest non-zero signal, corresponding to one Full-text assessment flag and three Exclude decisions. The overlaid line shows cumulative workload reduction if records below each score threshold were placed in the AI-excluded pool. The deployed process used the conservative rule that only 0% records were AI-excluded and all records scoring ≥12.5% were routed to human review.

**Extended Data Fig. 8.**
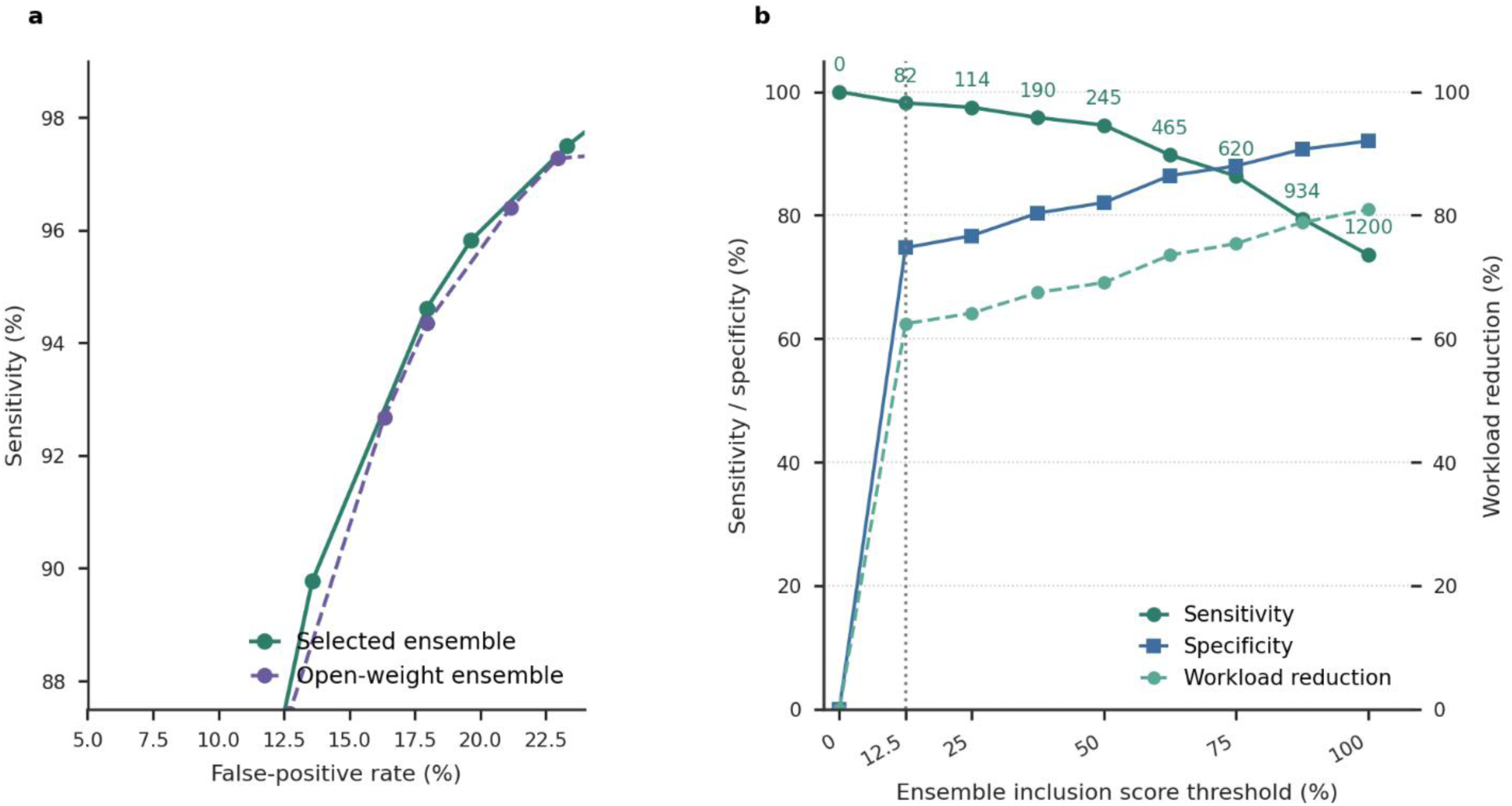
Ensemble score threshold trade-offs on 419-SR. a, ROC-style operating-point curve for the selected Gemini-3-flash plus GPT-5.1 ensemble and the open-weight Gemma-4-31B-IT plus GPT-OSS-120B ensemble, zoomed to the informative operating region. The selected operating point is highlighted. b, Sensitivity, specificity, and workload reduction as a function of ensemble score threshold. Solid lines show sensitivity and specificity for the selected ensemble; dashed lines show workload reduction for the selected ensemble. Numbers above the sensitivity curve indicate missed final-inclusion records at each threshold. The vertical dotted line marks the deployed operating rule: only records with a 0% ensemble score were AI-excluded; all records scoring ≥12.5% were routed to human review.

**Extended Data Table 1.** Performance of ten individual LLMs and the BioBERT comparator on 419 Cochrane reviews.

| Model | Family | Sensitivity (95% CI) | Specificity (95% CI) | TP | FN | TN | FP |
| --- | --- | --- | --- | --- | --- | --- | --- |
| GPT-OSS-120B | Open-weight (OpenAI) | 89.0% (87.0%-90.8%) | 87.8% (86.5%-89.0%) | 4079 | 466 | 18754 | 3593 |
| GPT-5 | OpenAI | 91.1% (89.4%-92.7%) | 89.6% (88.3%-90.8%) | 4140 | 405 | 19315 | 3032 |
| GPT-5.1 | OpenAI | 94.8% (93.4%-95.9%) | 86.5% (85.1%-87.9%) | 4293 | 252 | 18488 | 3859 |
| GPT-5.2 | OpenAI | 92.0% (90.4%-93.5%) | 87.9% (86.6%-89.2%) | 4203 | 342 | 18870 | 3477 |
| GLM-5 | Open-weight (Zhipu AI) | 93.1% (91.5%-94.6%) | 86.2% (84.6%-87.7%) | 4240 | 305 | 18223 | 4124 |
| Gemma-4-31B-IT | Open-weight (Google) | 95.2% (93.8%-96.4%) | 84.0% (82.4%-85.5%) | 4354 | 191 | 17729 | 4618 |
| Gemini-2.5-Pro | Gemini | 94.9% (93.6%-96.0%) | 83.7% (82.1%-85.2%) | 4337 | 208 | 17500 | 4847 |
| Gemini-3-Pro | Gemini | 95.6% (94.2%-96.7%) | 84.1% (82.4%-85.6%) | 4375 | 170 | 17734 | 4613 |
| Gemini-3.1-Pro | Gemini | 96.9% (95.9%-97.7%) | 83.9% (82.3%-85.5%) | 4396 | 149 | 17619 | 4728 |
| Gemini-3-flash | Gemini | 97.5% (96.6%-98.2%) | 81.5% (79.7%-83.4%) | 4432 | 113 | 16903 | 5444 |
| BioBERT (Chan et al.<sup>11</sup>) | Supervised | 97.5% | 65.0% | 4431 | 114 | 14516 | 7831 |

**Note:** Performance of ten individual LLMs, each evaluated using two independent runs with an OR rule, and the BioBERT comparator on 419-SR ( $n = 26,892$ records; 4,545 final-inclusion records). Sensitivity and specificity are review-level mean estimates with 95% review-clustered bootstrap percentile intervals; TP/FN/TN/FP are pooled record-level counts preserved for replication. BioBERT is retained as an external supervised baseline; review-level intervals are not shown for BioBERT because record-level predictions were not available from the comparator publication<sup>11</sup>. TP, true positive; FN, false negative; TN, true negative; FP, false positive. For the primary 419-SR endpoint, positives were records corresponding to final included studies.

## Supplementary information

## Supplementary notes

### Supplementary note 1 | Locked screening prompt

Two variants of the locked screening system prompt were used, matched to review context rather than to performance tuning. The extended retrospective variant was used for all Cochrane-derived analyses: 16-SR, 419-SR, the topic-matched external migraine and dementia validation subsets, and the 22-SR workload-estimation corpus. The concise prospective variant was used for live use inside the Swedish national migraine and dementia guideline programmes; it removed Cochrane-specific routing conventions that do not apply to national guideline reviews, where only completed evidence is screened, and tightened delivery modality and co-intervention clauses. The two variants share the same scaffold: role, three-class I/F/E decision, one-sentence reason, fixed three-column table output and the same user-prompt template. They differ only in the inclusion-convention clauses described below. Review-specific eligibility criteria were inserted into the user-prompt template: the review-level metadata fields released by Chan et al. (each review’s title, background, objectives and selection criteria) for 419-SR, 16-SR broad, the external migraine and dementia validation subsets, and 22-SR; or the full methods-section criteria for 16-SR detailed and prospective deployment. These general screening principles were subordinate to the review-specific eligibility criteria.

**Extended system prompt (retrospective analyses: 16-SR, 419-SR, external migraine and dementia validation, 22-SR)**

You are a systematic review expert. Does this study fulfill the criteria? Answer “include” or “Exclude”, if not sure answer “Fulltext assessment” and give the reason why in one sentence max. If study is a duplicate/from same study as one included before, then decide “Include” again. Make your answers in a numbered table. Column 1 = the number of the study, column 2 = inclusion/exclusion/full text as one letter I E or F, column 3 = inclusion/exclusion reason, one sentence max. Make your answers in a numbered table. Include study protocols, design/rationale papers and trial registry entries that describe an eligible study even if no results are yet reported; mark them as “ongoing/awaiting classification”. Delivery modality (in-person, virtual, hybrid) is not an exclusion criterion unless explicitly stated. If any arm tests the intervention alone against a suitable control, include the study even if other arms contain additional co-interventions. Subgroup or secondary analyses based on an eligible study are eligible when they report outcomes for the target population.

**Concise system prompt (prospective deployment: national migraine and dementia guidelines)**

You are a systematic review expert. Does this study fulfill the criteria? Answer include/exclude/fulltext as I/E/F with one short reason.

**User prompt template**

{criteria_text} CRITICAL INSTRUCTIONS: - The criteria above are authoritative and must be followed exactly. - Return a 3-column markdown table: | # | Decision | Reason | - Decision column: single letter only (I, E, or F). - Reason column: brief justification, one sentence max. --- STUDIES TO SCREEN --- {studies} --- END ({n} studies) --

### Supplementary note 2 | Dataset provenance, label definitions, and validation hierarchy

#### Label definitions in the Chan et al. dataset

The 419-SR and 22-SR datasets derive from the Chan et al. dataset and accompanying paper, whose authors searched on 15 December 2023 to collect data on systematic reviews published in the Cochrane Library; both are cited in the main manuscript.

419-SR dataset, approximate screening labels. Chan et al. used lists of included, excluded, and additional studies published with each Cochrane review to approximate the screening process. Each record receives a ternary label:

- Label 1.0 — Included studies: publications included in the final evidence synthesis.
- Label 0.5 — Excluded studies: publications that passed title and abstract screening but were excluded during full-text review.
- Label 0.0 — Additional studies: publications listed in the Cochrane review reference list that were neither finally included nor flagged as full-text excluded. The same definition applies to the external migraine and dementia validation subsets, which are drawn from the same Chan et al. partition.

As Chan et al. note, these labels only approximate the actual screening process. In particular, the additional study set (label 0.0) is much smaller than the set of publications actually excluded during real screening and remains related to the review topic because records appear in the review reference list. The 419-SR dataset therefore does not contain the large volume of clearly irrelevant citations encountered in a real literature search, and specificity estimates on 419-SR should be interpreted with this caveat.

22-SR dataset, real screening labels. In contrast, Chan et al. constructed 22-SR by replicating the actual database searches of 22 Cochrane reviews published between 15 December 2023 and 15 May 2024. This yielded real, unfiltered search results; label 0 records in 22-SR come from the replicated real search rather than from Cochrane reference-list approximation. The 22-SR dataset therefore provides a more realistic environment for workload-reduction estimation, with authentic label 0 records representing the spectrum of irrelevant citations encountered in real systematic review screening. The 22-SR LLM row uses Gemini-3-flash plus GPT-5, a near-predecessor workload-estimation pair, not the final Gemini-3-flash plus GPT-5.1 deployment pair

Prospective deployment labels. In the live migraine and dementia deployments, the analogous “label 0” class is the AI-excluded pool: records receiving Exclude (E) from all four AI runs. It is not a Cochrane reference-list construct; it is the pool not sent to first-pass human screening and subjected to prespecified audit.

The primary endpoint in the present study uses label = 1.0 (included studies) as the positive class. This is a deliberately conservative choice for the most consequential sensitivity question: in operational screening, the aim is to retain all studies that will ultimately be included, and missing a label-1.0 study constitutes a consequential error. Label 0.5 studies represent a harder intermediate category: records whose abstracts appeared relevant but whose full texts revealed ineligibility. Sensitivity at the broader label ≥0.5 threshold is reported in the supplementary performance tables.

Label 0.5 records are important for interpreting prospective title and abstract audit results. A title and abstract F decision does not necessarily imply final inclusion; it often represents a record worth inspecting at full text but ultimately excluded. Therefore, prospective AI-excluded records retained as F at blinded title and abstract consensus were treated as potential safety signals but not final false-negatives unless they remained finally retained or likely to change a review’s conclusions after consequence-based adjudication.

### Why multiple retrospective datasets were used

16-SR (development dataset) comprises 736 records from 16 Cochrane reviews published in May-June 2025 and therefore postdates the reported training cutoffs of the main retrospective models evaluated on 16-SR. In its raw reviewer-master datasets, label and label_corrected are binary: 1 = included after full-text assessment in the Cochrane authors’ EndNote libraries, 0 = excluded after full-text assessment; unlike Chan et al.-derived datasets, no 0.5 full-text-excluded category is used. Later-release GPT-5.5 models used in the criteria-richness sensitivity panel may partially overlap this publication window and are interpreted accordingly.

419-SR (primary validation) comprises 26,892 records from 419 reviews. Its scale enables precise confidence intervals for final inclusion sensitivity and supports subgroup analyses by review characteristics. The 419-SR dataset was drawn from the Chan et al. evaluation partition, distinct from the training partition used for the BioBERT model, enabling direct head-to-head comparison.

22-SR (workload-estimation) comprises 142,504 records from 22 replicated real searches. Unlike 419-SR, where label-0.0 records are additional studies drawn from Cochrane reference lists, 22-SR preserves raw, unfiltered search output, making it the dataset used for workload-reduction estimation. Its low prevalence (0.54% included records) provides the specificity estimate closest to routine search screening. After rehydration of the cached GPT-5 and Gemini-3-flash outputs, all 142,504 records have a populated selected-ensemble OR decision. The selected workload-estimation ensemble routed 13,968 records to human review, leaving the pooled workload reduction at 90.2%.

### Search dates and potential training data exposure

The Chan et al. 419-SR dataset was assembled from Cochrane reviews collected on 15 December 2023. The 22-SR dataset was constructed from reviews published between 15 December 2023 and 15 May 2024. Because LLM training corpora may include biomedical abstracts and Cochrane review metadata from this period, models could in principle recognise specific studies and inclusion labels rather than reason from eligibility criteria. Four findings make simple memorisation unlikely as the sole explanation: release series variability, stable disagreement phenotypes across datasets, temporal independence through 16-SR for the main retrospective models evaluated on that dataset, and prospective deployment on records retrieved after model training cutoffs. These checks do not exclude training data exposure.

### Supplementary note 3 | Kappa vigilance under class imbalance

An exploratory analysis tested whether lower agreement between two independent runs of the same model was associated with lower sensitivity at the review-level on 419-SR. Across ten models, model-level run-to-run κ versus OR-ensemble sensitivity showed Spearman ρ = 0.479 (p = 0.162). At the review-level for the selected ensemble components, the association was small but consistent: for Gemini-3-flash, ρ = 0.178 (p = 0.0007) across 361 reviews, with mean OR sensitivity 96.8% at or below the median κ and 98.2% above (mean missed inclusions 0.41 versus 0.21). For GPT-5.1, ρ was identical (0.178; p = 0.0007), with mean OR sensitivity 93.8% versus 95.7% (mean missed inclusions 0.92 versus 0.47). Lower within-model agreement was therefore associated with lower review-level sensitivity, but the effect was small. Agreement was therefore treated as a vigilance signal rather than an automatic stopping rule.

### Supplementary note 4 | Prospective migraine adjudication

A prespecified stratified random sample of 340 migraine records was adjudicated by two blinded human reviewers with consensus lock before AI outputs were unblinded. The sample included 300 AI-excluded records (E across all four runs) and 40 AI-flagged records.

No sampled AI-excluded record remained finally retained or likely to change a review’s conclusions after post-unblinding final adjudication, yielding 0 observed false-negatives in the prespecified AI-excluded stratum. Because final classification incorporated review of AI outputs and reasons, this endpoint evaluates the implemented AI-assisted screening and adjudication policy rather than a fully AI-independent diagnostic-accuracy reference standard.

A reconstructed blinded-consensus lock retained 17 AI-excluded records that were later reclassified as ineligible after AI outputs were unblinded, and excluded 2 AI-flagged records that were later revised to retain.

Using the uniform inverse-probability-weighted primary analysis specified for the prospective audit, migraine prospective sensitivity was estimated at 100.0% and specificity at 92.0% (bootstrap 95% CI, 89.8 94.5%). Sensitivity was identical under unweighted, uniform IPW, and design-weighted analyses because no final false-negatives were observed in the audited AI-excluded stratum.

Against the final post-unblinding standard, the reconstructed blinded lock had 22 true positives, 2 false-negatives and 17 false-positives, giving 91.7% sample sensitivity and 94.6% sample specificity.

Across the full adjudication sample, Reviewer A versus Reviewer B agreement was 82.6% raw agreement for three-category I/F/E decisions (Cohen’s κ = 0.32). Under screening-relevant binary retain-versus exclude coding (I/F versus E), raw agreement was 86.2% with κ = 0.44. In the AI-excluded stratum alone, binary raw agreement was 88.0% with κ = 0.22, illustrating the expected prevalence effect under a mostly negative stratum: kappa is suppressed by low positive prevalence even when raw agreement is high.

### Monitoring rule

The 340-record programme audits were interpreted using stepped escalation, drawing on the CESAR adaptive-platform protocol (cited in the main manuscript) as a reference for stepped harm monitoring rather than as an automatic stop rule. The primary safety denominator was the 300-record AI-excluded stratum within each programme. The per-programme audit was powered around a false-negative safety threshold rather than an expectation of zero false-negatives. With 300 audited records, <=8 false-negatives yields an upper one-sided exact binomial confidence bound below 5%. With 0 observed final false-negatives in each programme, the upper bound was approximately 1.0% per programme; pooled across 600 AI-excluded audited records, the one-sided exact upper bound was approximately 0.5%. Because the 419-SR selected ensemble had an observed unanimous-exclusion label-discordance rate below 1%, a small number of false-negatives would be compatible with benchmark performance; the prospective safety criterion instead tests whether the topic-local rate remains below the prespecified unacceptable threshold and whether any error is likely to change a review’s conclusions.

Major or likely to change a review’s conclusions false-negatives triggered review-specific containment regardless of the aggregate false-negative proportion. In the completed migraine and dementia audits, no final AI-exclusion error was observed, so no review-local or programme-level alarm was crossed.

Blinded-stage metrics reconstruct the pre-unblinding keep/exclude state using the documented post-unblinding revisions in the adjudication workbooks. This reconstruction is distinct from the main manuscript Table 2, which evaluates decision layers against the post-unblinding final consequence-based adjudicated standard. The separation is important because the final standard is appropriate for evaluating the implemented operational policy, but is not fully independent of the AI system.

### Supplementary note 5 | Prospective dementia adjudication

A prespecified stratified random sample of 340 dementia records used the same audit structure as the migraine programme: 300 records from the AI-excluded stratum and 40 from the AI-flagged stratum. Both strata have now been adjudicated against the original eligibility criteria. No sampled AI-excluded record remained finally retained after consequence-based adjudication. Two AI-excluded records that were retained at blinded consensus were reclassified as ineligible after AI outputs were unblinded. Among the 40 AI-flagged records, 14 were finally retained and 26 were finally excluded; five records initially excluded by locked blinded consensus were retained after AI outputs were unblinded. The final dementia AI-assisted confusion matrix in the audit sample was TP = 14, FP = 26, FN = 0 and TN = 300, giving 100.0% sample sensitivity and 92.0% sample specificity. The corresponding inverse-probability-weighted specificity was 90.6% (bootstrap 95% CI, 88.7-92.9). Reviewer A versus reviewer B agreement was 90.0% raw agreement with Cohen’s kappa = 0.23 for three-category decisions; binary retain versus exclude agreement was also 90.0% with kappa = 0.23.

At blinded consensus lock, 2 of 300 AI-excluded records were retained for further review and both were reclassified as ineligible after AI outputs were unblinded. In the AI-flagged stratum, 9 of 40 records were positive at lock and 5 additional records were revised from excluded to retained after AI unblinding, giving 14 final AI-flagged positives and 0 final AI-excluded positives (Supplementary Fig. 4).

## Supplementary methods

### Deduplication rules

Deduplication used rule-based exact matching after normalisation. Within PubMed, records were deduplicated by review group and PMID. Within EBSCO, records were deduplicated by search number and Study ID, or by title and year when no Study ID was available. In the combined PubMed–EBSCO dataset, records were deduplicated within review group using normalised DOI where available, otherwise normalised title and publication year, then title alone, and finally Study ID. Normalisation included case folding, whitespace normalisation, title cleanup, and DOI cleaning. PubMed records were preferentially retained over EBSCO records when duplicate records matched.

## Supplementary figures

**Supplementary Fig. 1.**
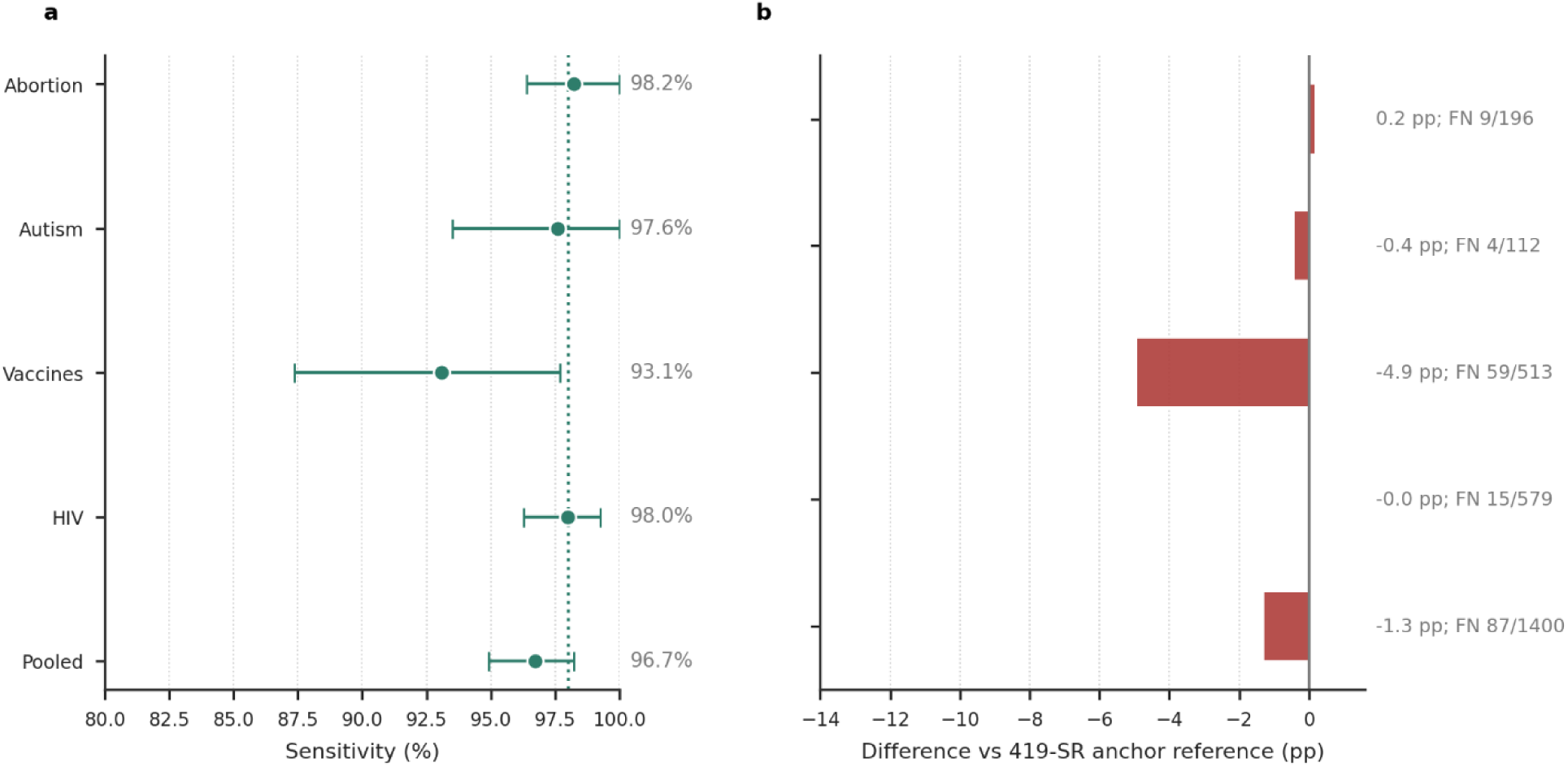
Normatively contested topic sensitivity on 419-SR matched comparator subsets. Selected LLM ensemble (Gemini-3-flash plus GPT-5.1) on politically sensitive or normatively contested topic subsets: abortion, autism, vaccines, and HIV, with the 419-SR anchor reference. a, Mean review-level sensitivity by topic; the dashed purple line marks the 419-SR anchor (review-level mean). b, Topic-specific difference from the 419-SR anchor in percentage points, with pooled record-level false-negative counts. These topics correspond to the heightened-oversight list prespecified in the operational protocol.

**Supplementary Fig. 2.**
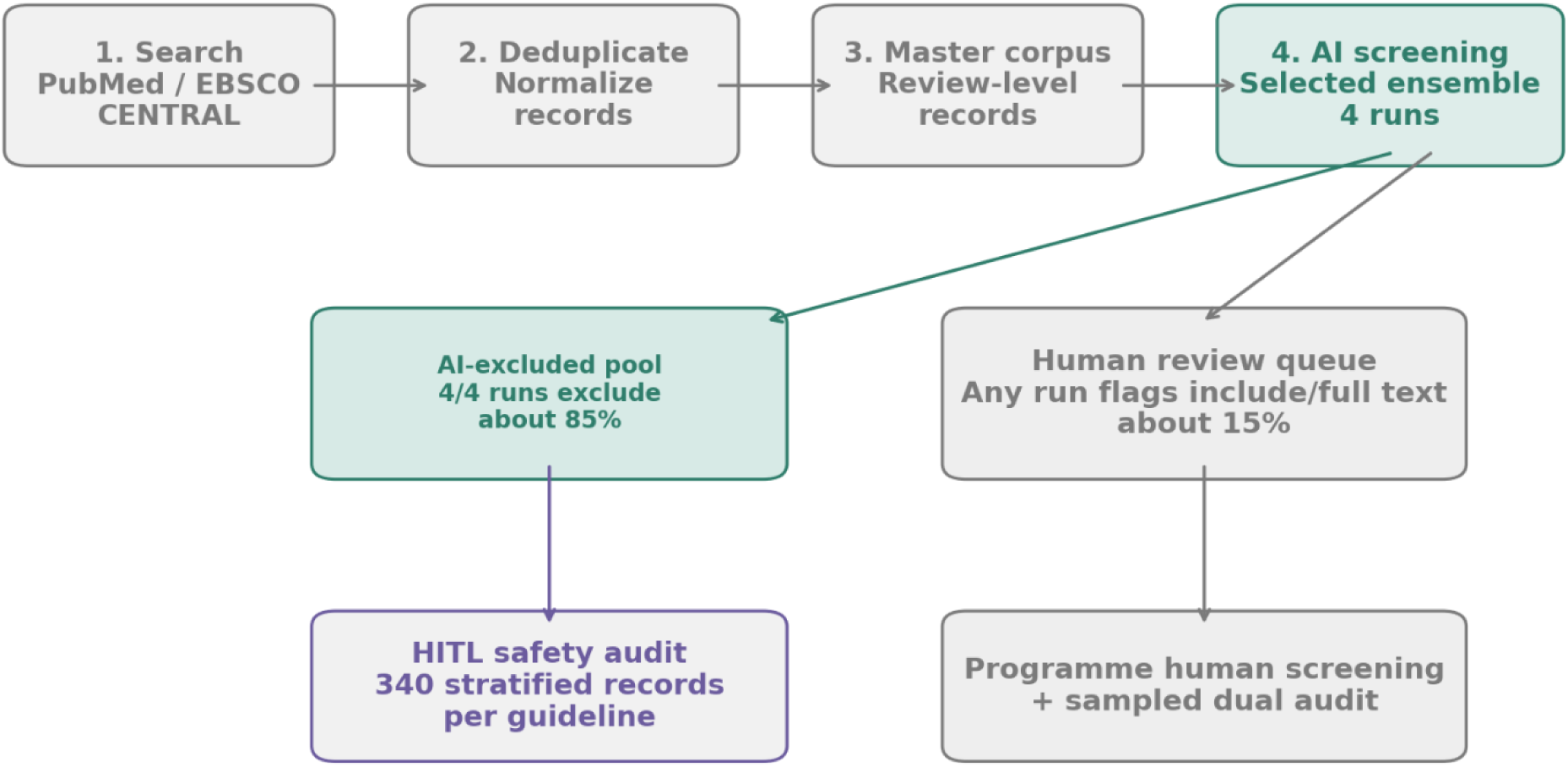
Automated search to screening pipeline. Programmatic search orchestration across PubMed, EBSCO databases, Cochrane CDSR and Cochrane CENTRAL; deduplication; assignment of stable study IDs; export to review-level workbooks; dual-model, dual-run AI screening with OR ensemble routing; and prespecified 340-record human audit comprising 300 AI-excluded and 40 AI-flagged records. The figure corresponds to STARD-AI item 24 (flow of records).

**Supplementary Fig. 3.**
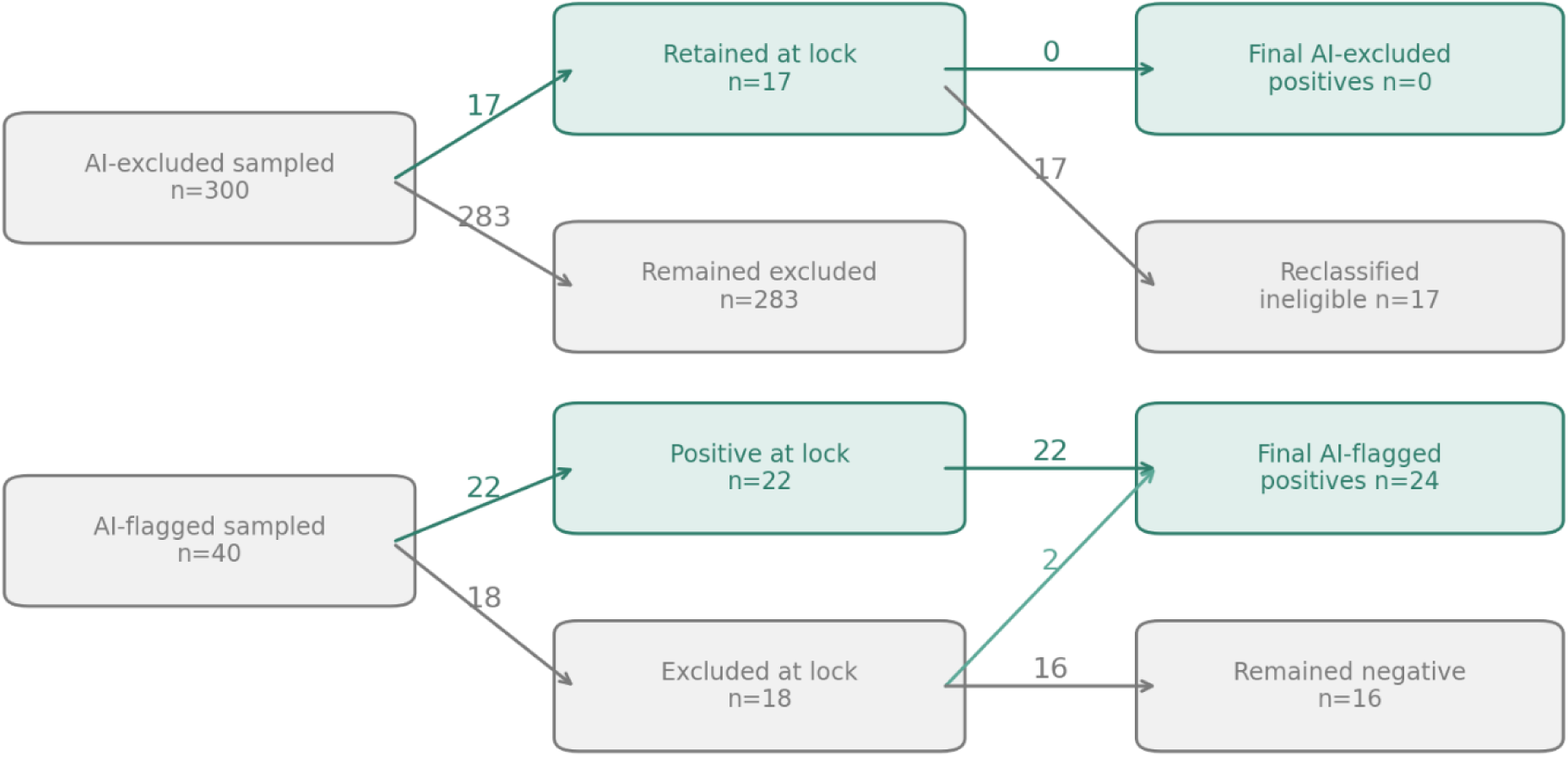
Prospective migraine adjudication flow. Flow of the prespecified 340-record migraine audit: 300 records sampled from the AI-excluded stratum (E across all four runs) and 40 from the AI-flagged stratum; dual blinded independent review; consensus lock; and secondary audit after AI outputs were unblinded. Final post-unblinding adjudication yielded 0 of 300 sampled AI-excluded false-negatives. The dementia programme used the same 340-record audit structure and is shown in Supplementary Fig. 4; completed numerical dementia results are retained in the prospective audit workbooks and summarised in main Table 2.

**Supplementary Fig. 4.**
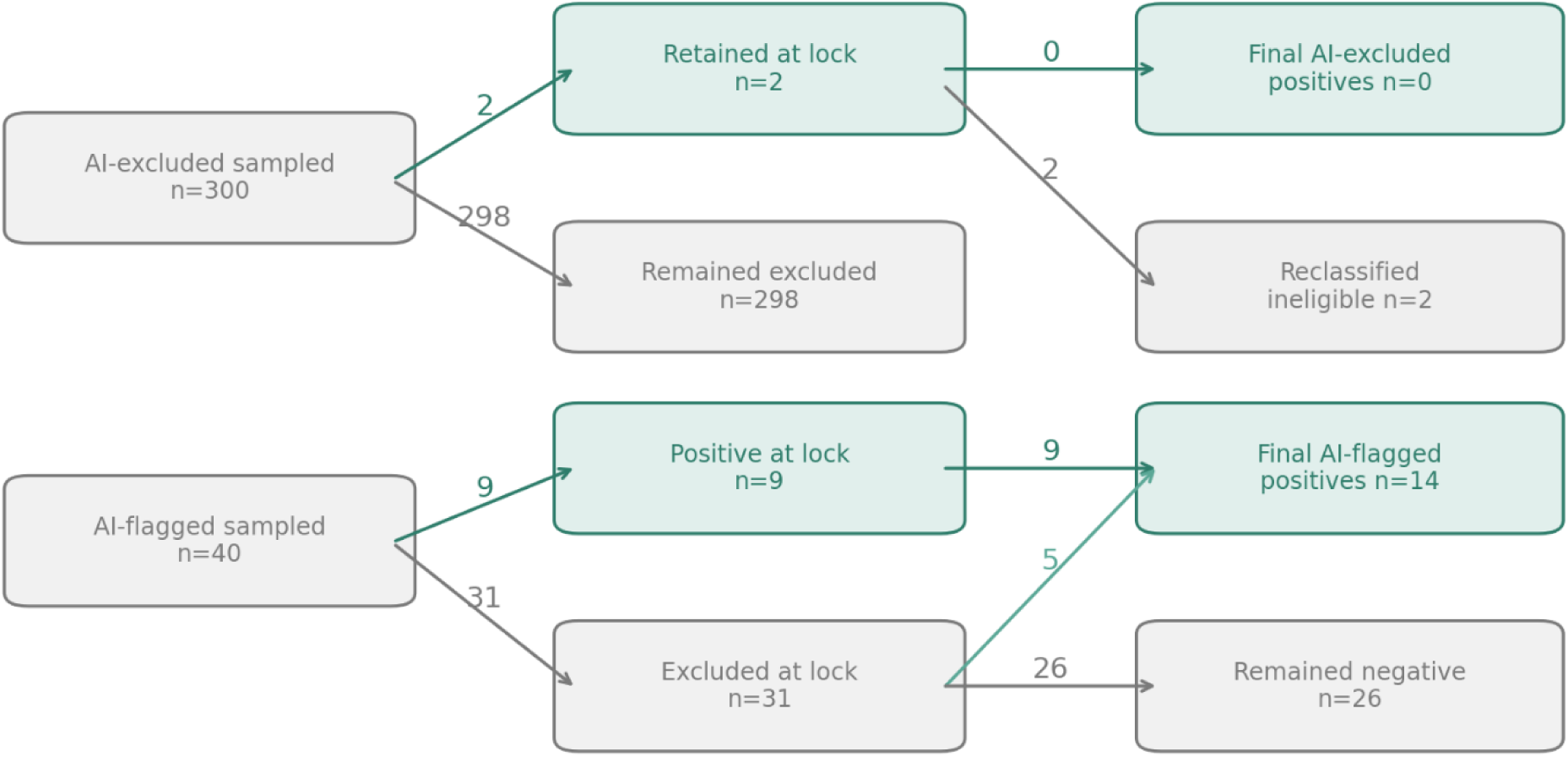
Prospective dementia adjudication flow. Flow of the prespecified 340-record dementia audit: 300 records sampled from the AI-excluded stratum (E across all four runs) and 40 from the AI-flagged stratum; dual blinded independent review; consensus lock before AI unblinding; and final adjudication after reviewing AI outputs and original review criteria. Two AI-excluded records retained at blinded lock were reclassified ineligible after unblinding, leaving 0 final AI-excluded positives under the post-unblinding adjudicated standard. Among AI-flagged records, 9 were positive at lock and 5 additional records were revised from excluded to retained, yielding 14 final AI-flagged positives.

## Supplementary tables and data tables

### Supplementary data files

Complete numerical tables are provided in the supplementary data tables. Supplementary Data Tables 6, 7 and 11 are not reproduced in full here because they are wider than an A4 page. The model card is also provided in machine-readable form; the brief version is embedded below.

**Supplementary Table 1.** Model specifications summary.

| Model | Family / access route | Role in package | Deployment type |
| --- | --- | --- | --- |
| Gemini-3-flash | Gemini Commercial API | Selected ensemble component | Dense / proprietary |
| GPT-5.1 | OpenAI Commercial API | Selected ensemble component | Dense / proprietary |
| Gemini-2.5-Pro | Gemini Commercial API | Comparator | Dense / proprietary |
| Gemini-3.1-Pro | Gemini Commercial API | Comparator | Dense / proprietary |
| GPT-5 | OpenAI Commercial API | Comparator and 22-SR workload-estimation ensemble component | Dense / proprietary |
| GPT-5.2 | OpenAI Commercial API | Comparator | Dense / proprietary |
| GPT-5.5 | OpenAI Commercial API | Comparator | Dense / proprietary |
| GLM-5 | Open-weight (Zhipu AI)<br>Vertex AI MaaS | Open-weight comparator | Open-weight (Apache-2.0) |
| GPT-OSS-120B | Open-weight (OpenAI)<br>Vertex AI MaaS | Open-weight comparator | Open-weight (Apache-2.0) |
| Gemma-4-31B-IT | Gemma (Google)<br>Vertex AI MaaS | Open-weight comparator | Open-weight (Apache-2.0) |

**Note:** Model families, access routes, roles in the package, and deployment types are summarised in this table and in the accompanying supplementary data tables. “Open-weight” denotes model weight availability, not necessarily local execution in this evaluation; open-weight models were evaluated through the specified managed inference route unless otherwise stated. Manuscript display names follow the package model registry; raw reviewer-master workbooks and cached-output columns may carry historical vendor or run identifiers (for example “gemini-3-flash preview” or “Gemini-3-flash-simple”) that map to the single display name “Gemini-3-flash” and do not denote separate models.

**Supplementary Table 2 | STARD-AI reporting checklist**

The STARD-AI reporting map is provided in the supplementary data tables and the reproducibility archive. Items 26a, 26b, 27, and 32 do not apply to citation-level screening because no patient-level diagnostic intervention was evaluated. All other items are Reported or Partially reported.

**Supplementary Table 3 | MI-CLAIM-GEN reporting checklist**

The MI-CLAIM-GEN checklist is provided in the supplementary data tables and the reproducibility archive. The checklist uses the MI-CLAIM-GEN item wording and maps each item to the manuscript or package location and study-specific note.

**Supplementary Table 4 | RAISE reporting guidance and responsible handover map for AI-assisted evidence synthesis screening**

The RAISE table is provided in the supplementary data tables and the reproducibility archive. It maps the study against the RAISE 2 reporting guidance and the RAISE 3 responsible handover tool, covering intended use, validation data, performance monitoring, user capability, risk assessment, licenses, transparency and reassessment triggers. RAISE guidance items are condensed into reader-facing reporting rows rather than reproduced verbatim.

Responsible handover. The evaluated tool is the Gemini-3-flash plus GPT-5.1 screening system: locked prompt, locked model versions, batch size, I/F/E output, two runs per model and OR-ensemble routing. It is intended for first-pass title and abstract triage in national guideline systematic reviews. Records may bypass first-pass human screening only when all four runs return E; any I or F signal sends the record to human review. Topic-local audit of the AI-excluded pool is required. Any change in model, prompt, provider, eligibility-criteria class or deployment context should be treated as a new version and re-evaluated before use.

**Supplementary Table 5.** Prospective deployment operational metrics.

| Metric | Migraine guideline | Dementia guideline | Combined |
| --- | --- | --- | --- |
| Systematic reviews | 16 | 8 | 24 |
| Records after deduplication | 36,503 | 38,176 | 74,679 |
| AI-excluded (E across all four runs) | 30,921 (84.7%) | 32,937 (86.3%) | 63,858 (85.5%) |
| Flagged for human review | 5,582 (15.3%) | 5,239 (13.7%) | 10,821 (14.5%) |
| Workload reduction | 84.7% | 86.3% | 85.5% |
| Prospective screening status | Complete | Complete | Complete |
| Human audit status | Complete | Complete | Complete |
| Locked adjudication sample | 340 | 340 | 680 |
| AI-excluded false-negative endpoint | 0/300 | 0/300 | 0/600 |
| Final retained records flagged by AI-assisted process | 24/24 | 14/14 | 38/38 |
| IPW sensitivity / specificity | 100.0% / 92.0% (89.8-94.5%) | 100.0% / 90.6% (88.7-92.9%) | 100.0% / 91.3% (89.8-93.1%) |

**Note:** Operational screening counts and adjudication outcomes are complete for both programmes. Prospective workload reduction (85.5% combined) is lower than the 22-SR real-search benchmark (90.2%) because the prospective corpus comprised structured guideline searches across six question categories with substantial type-targeted retrieval, yielding higher prevalence of records the AI ensemble routed for human review than the broader 22-SR Cochrane real-search distribution. Workload estimates in the main text use one minute per title and abstract decision and six screening hours per reviewer-day; at 30 seconds, one minute and two minutes per record, conventional dual screening would require approximately 207, 415 and 830 person-days, while the AI-assisted process would require approximately 17, 34 and 68 person-days before implementation, audit-meeting and maintenance overheads.

**Supplementary Data Table 6 | Prospective migraine guideline per-review AI screening statistics**

The complete numerical table is provided as the corresponding Supplementary Data Table. It reports review identifier, topic type, record count, LLM ensemble flagged records, AI-excluded records, workload reduction, Gemini-only and GPT-only flag counts, per-run positive call counts, raw agreement, Cohen’s κ within model families, and between-family agreement. Note: Raw agreement and per-run positive-call counts are shown alongside Cohen’s κ because, under extreme class imbalance, κ can be low even when raw agreement is very high. Workload reduction is calculated as AI-excluded records divided by all deduplicated records in the review.

**Supplementary Data Table 7 | Prospective dementia guideline per-review AI screening statistics**

The complete numerical table is provided as the corresponding Supplementary Data Table. It uses the same column structure as Supplementary Data Table 6 for the dementia guideline reviews, including review identifier, topic type, record count, LLM ensemble flagged records, AI-excluded records, workload reduction, per-run positive-call counts and agreement summaries.

**Supplementary Table 8.**
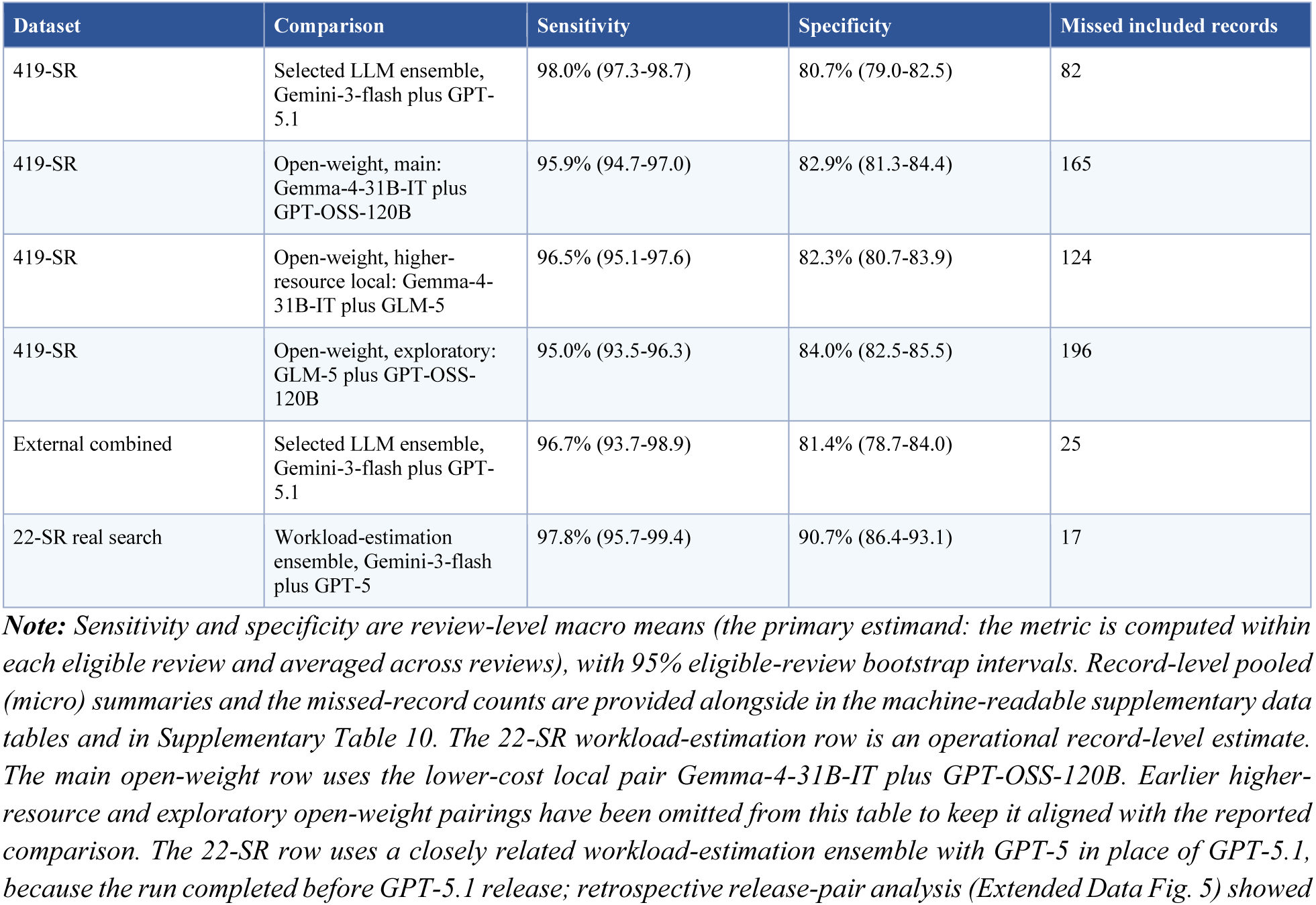
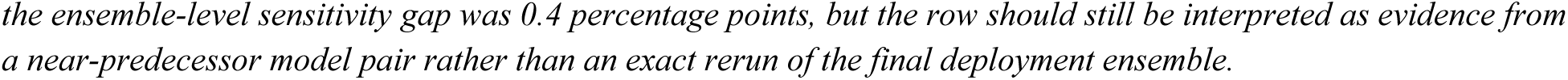
Open-weight versus selected and workload estimation LLM ensembles across packaged datasets.

| Dataset | Comparison | Sensitivity | Specificity | Missed included records |
| --- | --- | --- | --- | --- |
| 419-SR | Selected LLM ensemble, Gemini-3-flash plus GPT-5.1 | 98.0% (97.3-98.7) | 80.7% (79.0-82.5) | 82 |
| 419-SR | Open-weight, main: Gemma-4-31B-IT plus GPT-OSS-120B | 95.9% (94.7-97.0) | 82.9% (81.3-84.4) | 165 |
| 419-SR | Open-weight, higher-resource local: Gemma-4-31B-IT plus GLM-5 | 96.5% (95.1-97.6) | 82.3% (80.7-83.9) | 124 |
| 419-SR | Open-weight, exploratory: GLM-5 plus GPT-OSS-120B | 95.0% (93.5-96.3) | 84.0% (82.5-85.5) | 196 |
| External combined | Selected LLM ensemble, Gemini-3-flash plus GPT-5.1 | 96.7% (93.7-98.9) | 81.4% (78.7-84.0) | 25 |
| 22-SR real search | Workload-estimation ensemble, Gemini-3-flash plus GPT-5 | 97.8% (95.7-99.4) | 90.7% (86.4-93.1) | 17 |

**Note:** Sensitivity and specificity are review-level macro means (the primary estimand: the metric is computed within each eligible review and averaged across reviews), with 95% eligible-review bootstrap intervals. Record-level pooled (micro) summaries and the missed-record counts are provided alongside in the machine-readable supplementary data tables and in Supplementary Table 10. The 22-SR workload-estimation row is an operational record-level estimate. The main open-weight row uses the lower-cost local pair Gemma-4-31B-IT plus GPT-OSS-120B. Earlier higher-resource and exploratory open-weight pairings have been omitted from this table to keep it aligned with the reported comparison. The 22-SR row uses a closely related workload-estimation ensemble with GPT-5 in place of GPT-5.1, because the run completed before GPT-5.1 release; retrospective release-pair analysis (Extended Data Fig. 5) showed
the ensemble-level sensitivity gap was 0.4 percentage points, but the row should still be interpreted as evidence from a near-predecessor model pair rather than an exact rerun of the final deployment ensemble.

**Supplementary Table 9 | Cochrane Review Groups represented in the 419-SR validation dataset**

Acute Respiratory Infections (ARI); Airways (AIRWAYS); Anaesthesia (ANAESTH); Back and Neck (BACK); Bone, Joint and Muscle Trauma (MUSKINJ); Breast Cancer (BREASTCA); Childhood Cancer (CHILDCA); Colorectal (COLORECT); Common Mental Disorders (DEPRESSN); Consumers and Communication (COMMUN); Cystic Fibrosis and Genetic Disorders (CF); Dementia and Cognitive Improvement (DEMENTIA); Developmental, Psychosocial and Learning Problems (BEHAV); Drugs and Alcohol (ADDICTN); Effective Practice and Organisation of Care (EPOC); Emergency and Critical Care (ECC); ENT; Epilepsy; Eyes and Vision (EYES); Fertility Regulation (FERTILREG); Gut; Gynaecological, Neuro-oncology and Orphan Cancer (GYNAECA); Gynaecology and Fertility (MENSTR); Haematology (HAEMATOL); Heart; Hepato-Biliary (HEPATO); HIV; Hypertension (HTN); Incontinence (INCONT); Infectious Diseases (INFECTN); Injuries (INJ); Kidney and Transplant (RENAL); Lung Cancer (LUNGCA); Metabolic and Endocrine Disorders (ENDOC); Methodology (METHOD); Movement Disorders (MOVEMNT); Multiple Sclerosis and Rare Diseases of the CNS (MS); Musculoskeletal (MUSKEL); Neonatal; Neuromuscular (NEUROMUSC); Oral Health (ORAL); Pain, Palliative and Supportive Care (SYMPT); Pregnancy and Childbirth (PREG); Public Health (PUBHLTH); Schizophrenia (SCHIZ); Sexually Transmitted Infections (STI); Skin; Stroke; Tobacco Addiction (TOBACCO); Urology (UROL); Vascular (PVD); Work (OCCHEALTH); Wounds.

**Supplementary Table 10.** Micro and macro sensitivity summaries.

| Dataset | N reviews | N reviews with<br>≥1 included | N records | N included<br>records | Macro mean<br>sensitivity<br>(primary) | Macro median<br>sensitivity | Micro<br>sensitivity<br>(secondary) | Zero missed |
| --- | --- | --- | --- | --- | --- | --- | --- | --- |
| 419-SR selected ensemble | 419 | 361 | 26,892 | 4,545 | 98.0% (97.3-98.7) | 100.0% (IQR 100-100) | 98.2% | 305 |
| 419-SR open-weight ensemble | 419 | 361 | 26,892 | 4,545 | 95.9% (94.7-97.0) | 100.0% (IQR 98.1-100) | 96.4% | 268 |
| 22-SR workload estimation | 22 | 22 | 142,504 | 768 | 98.3% (96.8-99.5) | 100.0% (IQR 97.7-100) | 97.8% | 16 |
| External migraine | 27 | 25 | 1,343 | 293 | 99.1% (98.0-100) | 100.0% (IQR 100-100) | 98.6% | 22 |
| External dementia | 106 | 87 | 7,158 | 671 | 96.0% (92.3-98.8) | 100.0% (IQR 100-100) | 96.9% | 74 |
| External combined | 133 | 112 | 8,501 | 964 | 96.7% (93.7-98.9) | 100.0% (IQR 100-100) | 97.4% | 96 |

**Note:** Macro mean review-level sensitivity is the primary inferential estimand reported throughout the manuscript; pooled record-level micro sensitivity is reported as a secondary descriptive summary to show how review clustering changes the summary. Primary CIs are review-level bootstrap percentile intervals ( $B = 2,000$ , seed = 42) in which, for each estimand, only the eligible reviews are resampled—reviews with at least one included record for sensitivity and reviews with at least one non-included record for specificity.

**Supplementary Data Table 11 | 16-SR broad versus detailed drift across GPT and Gemini models**

The complete numerical table is provided as Supplementary Data Table 11. Performance is evaluated against corrected labels. Confidence intervals are review-clustered bootstrap 95% intervals. The table has been limited to submitted criteria-richness models and excludes development-only prompt-tuning rows and older 16-SR-only comparators. Gemini-3-Pro was not evaluated under detailed criteria because the model was deprecated from Google API service as of April 2026.

**Supplementary Table 12.** Selected versus higher-cost frontier ensemble on 16-SR.

| Criteria | Ensemble | Sensitivity | Specificity | Missed | Cost USD/1k |
| --- | --- | --- | --- | --- | --- |
| Broad | GPT-5.5 plus Gemini-3.1-Pro | 94.5% (85.7-100.0) | 46.2% (28.8-68.2) | 18 | Not measured |
| Broad | Gemini-3-flash plus GPT-5.1 | 100.0% (100.0-100.0) | 40.0% (24.1-60.0) | 0 | Not measured |
| Detailed | GPT-5.5 plus Gemini-3.1-Pro | 100.0% (100.0-100.0) | 51.8% (34.0-73.9) | 0 | 13.61 |
| Detailed | Gemini-3-flash plus GPT-5.1 | 99.7% (99.3-100.0) | 42.5% (24.9-67.8) | 1 | 5.46 |

**Note:** Performance evaluated against corrected labels. Confidence intervals are review-clustered bootstrap 95% intervals. Costs are measured only for detailed-criteria runs and reported as total cost per 1,000 records. Criteria specification changed the selected ensemble's missed-record set on 16-SR: broad criteria yielded 329 of 329 sensitivity (0 missed retained records), whereas detailed criteria yielded 328 of 329 (1 missed); specificity also differed.

**Supplementary Table 13.** Post hoc release-pair ensemble drift on 419-SR.

| Ensemble | Sensitivity macro primary 95% CI | Specificity macro primary 95% CI | Sensitivity micro secondary | Specificity micro secondary | AI-excluded pool | Routed to human | Discord. % |
| --- | --- | --- | --- | --- | --- | --- | --- |
| Gemini-2.5-Pro + GPT-5 | 95.79 (94.5-96.8) | 82.98 (81.4-84.6) | 96.30 | 77.55 | 17,497 | 9,395 | 0.96 |
| Gemini-3-Pro + GPT-5.1 | 97.21 (96.2-98.0) | 82.37 (80.7-84.0) | 97.51 | 77.17 | 17,359 | 9,533 | 0.65 |
| Gemini-3.1-Pro + GPT-5.2 | 97.28 (96.4-98.0) | 83.13 (81.5-84.8) | 97.25 | 77.83 | 17,518 | 9,374 | 0.71 |
| Selected: Gemini-3-flash + GPT-5.1 | 98.03 (97.3-98.7) | 80.75 (79.0-82.5) | 98.20 | 74.73 | 16,783 | 10,109 | 0.49 |

**Note:** Post hoc comparison of temporally ordered dual-model, dual-run release-pair ensembles and the selected frozen deployment ensemble. Sensitivity uses label 1.0 as positive; labels 0.0 and 0.5 are negative. Records were routed to human review if any of the four runs returned I or F; only records receiving E across all four runs were placed in the AI-excluded pool. The macro (primary) columns are review-level macro means with review-clustered bootstrap 95% percentile intervals ( $B = 2,000$ , seed = 42). The micro (secondary) columns are pooled record-level summaries from the same TP/FN/TN/FP and are reported for traceability with prior versions of this table; they are not the primary inferential estimand. The selected frozen ensemble row (highlighted) is the same Gemini-3-flash + GPT-5.1 pair used in external validation and prospective deployment. Because 419-SR labels were not re-adjudicated after AI review, the discordance rate should be interpreted as an observed AI-label discordance rate and may overestimate confirmed false-negative exclusions.

**Supplementary Table 14.** Audit monitoring and stepped escalation rules.

| Monitoring domain | Trigger | Level | Action |
| --- | --- | --- | --- |
| Inconsequential difference | Disagreement in rationale or caution only | Green | Record and continue |
| One moderate AI exclusion error | One eligible missed record judged unlikely to change review conclusions | Yellow | Expand audit in affected review |
| One major AI exclusion error | One eligible missed record judged plausibly review-changing | Orange | Immediate review-specific containment; expand audit; route similar records to human review |
| Repeated review-local errors | Additional major or repeated moderate errors in same review | Orange/Red local | Senior methodological review of affected review; potential suspension of AI exclusion for that review |
| Cross-review mechanism | Same error phenotype in $\geq 2$ reviews | Red programme | Senior methodological review; potential suspension of affected record class or programme process pending review |
| Absolute harm ceiling | Major-error point estimate $>3\%$ | Red programme | Senior methodological review; potential suspension of AI exclusion across programme |
| Statistical harm ceiling | Lower 95% exact CI bound for major-error proportion $>2\%$ | Red programme | Senior methodological review; potential suspension of AI exclusion across programme |
| Human-comparator non-inferiority | AI sensitivity below locked human consensus sensitivity by $\geq 5$ percentage points | Red programme | Senior methodological review; potential process suspension or redesign |
| Human-comparator false exclusion | Single-human reviewer or locked consensus excludes a final retained record | Comparator context | Record; include in reviewer versus AI comparison; assess repeated phenotype |
| Human-comparator workload error | Single-human reviewer or locked consensus retains/routes final excluded records | Workload context | Record; include in specificity/workload interpretation |
| Technical failure | Prompt/model/version/parsing/data-processing fault | Red programme | Suspend until corrected and revalidated |
| Unanimous-exclusion FN safety criterion | $>8$ FNs among 300 audited AI-excluded records | Red | Senior methodological review; pause routine AI exclusion; expand audit; potential redesign or suspension |
| Conclusion-changing AI-exclusion miss | Any likely to change a review's conclusions missed record | Orange local | Immediate containment and expanded audit regardless of aggregate rate |

**Supplementary Table 15 | Model card for the evaluated LLM screening system**

The model card is embedded as Supplementary Table 15 and also provided in machine-readable form in the supplementary data tables. It summarises intended use, out-of-scope use, model components, inputs, outputs, validation evidence, human oversight, known limitations, reassessment triggers and transparency package contents.

**Supplementary Table 16.** Per-dataset mapping of eligibility criteria source and system prompt variant.

| Dataset | Eligibility criteria fed to the prompt | System prompt variant |
| --- | --- | --- |
| 16-SR (broad) | Chan et al.-style review-level metadata (title, background, objectives, selection criteria) | Extended |
| 16-SR (detailed) | Full methods-section criteria | Extended |
| 419-SR | Chan et al. review-level metadata (title, background, objectives, selection criteria) | Extended |
| External migraine validation | Chan et al. review-level metadata (title, background, objectives, selection criteria) | Extended |
| External dementia validation | Chan et al. review-level metadata (title, background, objectives, selection criteria) | Extended |
| 22-SR | Chan et al. review-level metadata (title, background, objectives, selection criteria) | Extended |
| Prospective migraine | Full methods-section criteria | Concise |
| Prospective dementia | Full methods-section criteria | Concise |

**Note.** The extended variant retains Cochrane-specific routing conventions appropriate to Cochrane-derived analyses, including study protocols, design or rationale papers, and trial-registry entries describing eligible studies classed as “ongoing/awaiting classification”. The concise variant removes these clauses because Swedish national guideline reviews exclude protocols and ongoing studies and operate on completed evidence only. The user-prompt template was identical across variants. The difference between broad and detailed criteria was tested on 16-SR and is reported in Extended Data Fig. 2.

## Notes

### Competing Interest Statement

The authors have declared no competing interest.

